# Two schizophrenia imaging signatures and their associations with cognition, psychopathology, and genetics in the general population

**DOI:** 10.1101/2022.01.07.22268854

**Authors:** Ganesh B. Chand, Pankhuri Singhal, Dominic B. Dwyer, Junhao Wen, Guray Erus, Jimit Doshi, Dhivya Srinivasan, Elizabeth Mamourian, Erdem Varol, Aristeidis Sotiras, Gyujoon Hwang, Paola Dazzan, Rene S. Kahn, Hugo G. Schnack, Marcus V. Zanetti, Eva Meisenzahl, Geraldo F. Busatto, Benedicto Crespo-Facorro, Christos Pantelis, Stephen J. Wood, Chuanjun Zhuo, Russell T. Shinohara, Haochang Shou, Yong Fan, Nikolaos Koutsouleris, Antonia N. Kaczkurkin, Tyler M. Moore, Anurag Verma, Monica E. Calkins, Raquel E. Gur, Ruben C. Gur, Marylyn D. Ritchie, Theodore D. Satterthwaite, Daniel H. Wolf, Christos Davatzikos

## Abstract

The prevalence and significance of schizophrenia-related phenotypes at the population-level are debated in the literature. Here we assess whether two recently reported neuroanatomical signatures of schizophrenia, signature 1 with widespread reduction of gray matter volume, and signature 2 with increased striatal volume, could be replicated in an independent schizophrenia sample, and investigate whether expression of these signatures can be detected at the population-level and how they relate to cognition, psychosis spectrum symptoms, and schizophrenia genetic risk. This cross-sectional study used an independent schizophrenia-control sample (n=347; age 16-57 years) for replication of imaging signatures, and then examined two independent population-level datasets: Philadelphia Neurodevelopmental Cohort [PNC; n=359 typically developing (TD) and psychosis-spectrum symptoms (PS) youth] and UK Biobank (UKBB; n=836; age 44-50 years) adults. We quantified signature expression using support-vector machine learning, and compared cognition, psychopathology, and polygenic risk between signatures. Two neuroanatomical signatures of schizophrenia were replicated. Signature 1 but not signature 2 was significantly more common in youth with PS than TD youth, whereas signature 2 frequency was similar. In both youth and adults, signature 1 had worse cognitive performance than signature 2. Compared to adults with neither signature, adults expressing signature 1 had elevated schizophrenia polygenic risk scores, but this was not seen for signature 2. We successfully replicate two neuroanatomical signatures of schizophrenia, and describe their prevalence in population-based samples of youth and adults. We further demonstrate distinct relationships of these signatures with psychosis symptoms, cognition, and genetic risk, potentially reflecting underlying neurobiological vulnerability.

## INTRODUCTION

Schizophrenia (SZ) is a neuropsychiatric disorder imposing a significant personal and socioeconomic burden (1). Understanding the neuropathological processes underlying SZ has been hampered by the neurobiological heterogeneity of this disorder (2, 3), which has confounded case-control studies and impeded progress in treatment development (4). There is a pressing need for quantitative phenotypes and precision diagnostics, which could help define optimal personalized intervention according to a patient’s neurobiological and clinical profile. Machine learning applied to neuroimaging has emerged as a valuable tool to establish precise and quantitative imaging phenotypes. Specifically, our recent study from the PHENOM consortium revealed the presence of two statistically optimal and distinct neuroanatomical signatures in SZ (5). The first was characterized by widespread reductions of white matter (WM) and gray matter (GM) volume correlating with illness duration and worse premorbid functioning, while the second was associated with overall normal brain structure except for enlarged striatal and internal capsule volumes. This suggests two different underlying neuropathological mechanisms and highlights the need to both replicate these signatures in independent schizophrenia samples, and to investigate these signatures in population-based samples without schizophrenia.

Intense effort is now focused on understanding the pathophysiology of early stages of psychosis (6, 7) which will be essential for prevention and early intervention, and has the additional advantage of avoiding chronic effects of illness or treatment. Most psychosis-risk research examines “clinical high risk” groups, which are typically help-seeking and identified in clinical settings (8). However, longstanding theories of schizotaxia (biological vulnerability to SZ) and schizotypy (the wide spectrum of psychological manifestations of that vulnerability) argue that biological vulnerability to SZ is expressed in a substantial proportion of the general population, many of whom will never develop psychosis (9, 10). Examination of SZ-related neurostructural phenotypes in population-based samples is therefore critical to lay the groundwork for primary prevention efforts that incorporate stratification based on critical factors driving heterogeneity (6, 7). Prior work in population-based samples has demonstrated that psychosis spectrum symptoms (PS) are associated with increased risk of psychotic conversion (∼10% convert) (11, 12). SZ-associated structural brain abnormalities are also linked to PS in population samples (13). Whether the two SZ imaging signature expressions (“signature 1” and “signature 2”) are detectable in population-based PS is unknown. Even in healthy populations, SZ genetic risk is associated with regional brain differences resembling signature 1 (fronto-temporal reductions) or signature 2 (basal ganglia enlargement) (14, 15); however, whether these signatures associate with schizophrenia polygenic risk scores (PRS_SZ_) in population-based samples is unknown. To address these gaps, the present study replicated the signatures in schizophrenia, and then leveraged two independent population-level (non-help-seeking) cohorts. First, the Philadelphia Neurodevelopmental Cohort (PNC) (13), which offers rich cognition and psychopathology data in PS and typically developing (TD) youths, was analyzed to investigate the presence of signature 1 and signature 2 expressions and their clinical and cognitive correlates. Second, the UK Biobank (UKBB) (16), which offers limited cognition and rich genetics data in adults, was analyzed to explore the prevalence of these signatures and their cognitive and genetic correlates.

We hypothesized that SZ neuroanatomical signatures would be replicated in independent schizophrenia samples, and would be more common and prominent in the psychosis-spectrum group from the non-help-seeking PNC youth sample. We expected signature 1 but not signature 2 to be associated with poorer cognition, based on the known relationship between reduced gray matter volume (observed only in signature 1) and worse cognition (17-19). Given that PRS_SZ_ was defined using a large SZ sample encompassing all potential SZ subtypes (20, 21) we hypothesized that both signatures would show elevated PRS_SZ_. However, we hypothesized this elevation would be stronger in signature 1 than signature 2, because PRS_SZ_ is most strongly elevated in schizophrenia associated with early neurodevelopmental insults (22), and is also associated with fronto-temporal cortical reductions (14) and cognitive impairment (23), and because we observed signature 1 to be more prevalent than signature 2 in schizophrenia, likely influencing the allelic composition of the PRS_SZ._

## MATERIALS AND METHODS

### Study sample

This study includes human participants (n = 2,213) from the PHENOM consortium (n = 671), replication data (n = 347), PNC (n = 359 TD and PS), and UKBB (n = 836). The PHENOM sample includes participants with established SZ (n = 307) and healthy controls (n = 364). PNC is a large-scale community-based study, and after restricting age 16 years or older (to ensure overlapping age with the PHENOM sample so that machine learning models can be applied appropriately), there were 181 PS and 178 TD individuals. The UKBB is a large-scale open access resource for neuroimaging, genetic, and limited cognitive datasets. The subsample of UKBB healthy participants age 50 years or below were analyzed. Samples are detailed in the **supplement**.

### Image acquisition and preprocessing

A **mu**lti-atlas region **s**egmentation utilizing **e**nsembles of registration algorithms and parameters, and locally optimal atlas selection (MUSE) (24) was used to segment each individual’s T1-weighted images into 145 anatomical regions of interest (ROIs) from GM, WM, and cerebrospinal fluid **(Table S1)**. Voxel-wise volumetric maps for GM and WM were generated by deformable registration of skull-stripped T1-images to MNI space (see **supplement**).

### Identifying the presence of SZ imaging signatures and defining subgroups

**H**eterogeneit**y** through **d**isc**r**iminative **a**nalysis (HYDRA) (25) was used to identify the presence and expression strength of SZ imaging signatures. In contrast to unsupervised or fully-supervised clustering techniques, HYDRA is a semi-supervised machine learning method and uses pre-specified patient and control labels but then uses a data-driven approach to simultaneously perform classification and clustering *within* the patient group. Rather than forcing patient data into a single common discriminative pattern, HYDRA allows for the separation of distinct patient groups. The HYDRA parameters derived from PHENOM (5) were applied to harmonized PNC and UKBB data using support-vector machine learning (25, 26) to estimate neuroanatomical signature expression strength (E_1_ and E_2_). Since controls are assigned ‘-1’ and SZ ‘+1’ during HYDRA training, E (> 0) represents signature presence while E (< 0) represents its absence. Thus, each participant was assigned to one of these subgroups: signature 1 (E_1_ > 0, E_2_ < 0), signature 2 (E_1_ < 0, E_2_ > 0), none of signatures (E_1_ < 0, E_2_ < 0), and both signatures (E_1_ > 0, E_2_ > 0) (see **supplement**). These subgroups were compared for clinical, cognitive, genetic, and voxel-wise analyses. Signature expression strengths were also compared between TD and PS in PNC, and healthy controls vs. SZ in PHENOM.

### Voxel-wise volumetric analyses

To visualize voxel-wise volumetric differences between signatures, we used **m**ult**i**variate **d**iscrimin**a**tive **s**tatistical mapping (MIDAS) (27). MIDAS determines the optimal regional smoothing and provides higher sensitivity and specificity in identifying group alterations compared to other methods (see **supplement**).

### Clinical and cognitive measures

To assess clinical profiles, we used psychopathology scores derived from the PNC structured interview (28). To evaluate PNC cognition, we utilized the Penn Computerized Neurocognitive Battery (CNB), analyzing summary z-score measures of factors capturing overall accuracy, speed, and efficiency (29). Primary analyses of cognition in UKBB were limited to the fluid intelligence (30) and trail making test part-B (TMT-B) (31) based on their availability in larger samples (see further details in **supplement ‘**Study samples’ and ‘Clinical measures’).

### Genetic measures

Since the PRS_SZ_ is only valid for the European-ancestry population used to derive it (32), we limited PRS_SZ_ calculation to the UKBB European-ancestry cohort (n = 671) from the n = 836 full sample. We used validated PRS_SZ_ weights from the Polygenic Score Catalog (20) generated from 284,262 SNPs (21). LiftOver was used to convert SNPs to build GRCh38 (33), and PLINK 1.9 was used to calculate PRS_SZ_ (34).

### Statistical analyses

The proportions of participants expressing each of the signatures were compared between groups using a chi-square test. A two-sided Wilcoxon rank sum test compared cognitive, clinical, and PRS measures. Spearman correlations were used to examine associations between dimensional signature expression strength and cognition or PRS. We applied non-parametric tests across comparisons because some measures, especially TMT-B, were not normally distributed even after applying transformation. Age and sex covariates were controlled in all analyses. Ancestry principal components were also controlled in genetic analyses. P-values were corrected for multiple comparisons using false-discovery rate (FDR), requiring FDR-corrected p < 0.05.

## RESULTS

### Replication of SZ neuroanatomical subtypes

SZ neuroanatomical subtyping using HYDRA replicated the same two subtype profiles in the independent SZ sample (**Figure S1-S2**), with proportions (58.39% signature 1, 41.61% signature 2) similar to those in the original PHENOM sample.

### Prevalence of SZ imaging signatures in the PNC

In TD, prevalence of SZ signature 1, signature 2, both signatures, and none of SZ signatures (‘None’) was 23.03%, 24.16%, 5.62%, and 47.19%, respectively (**Figure 1A**).

**Figure 1.**
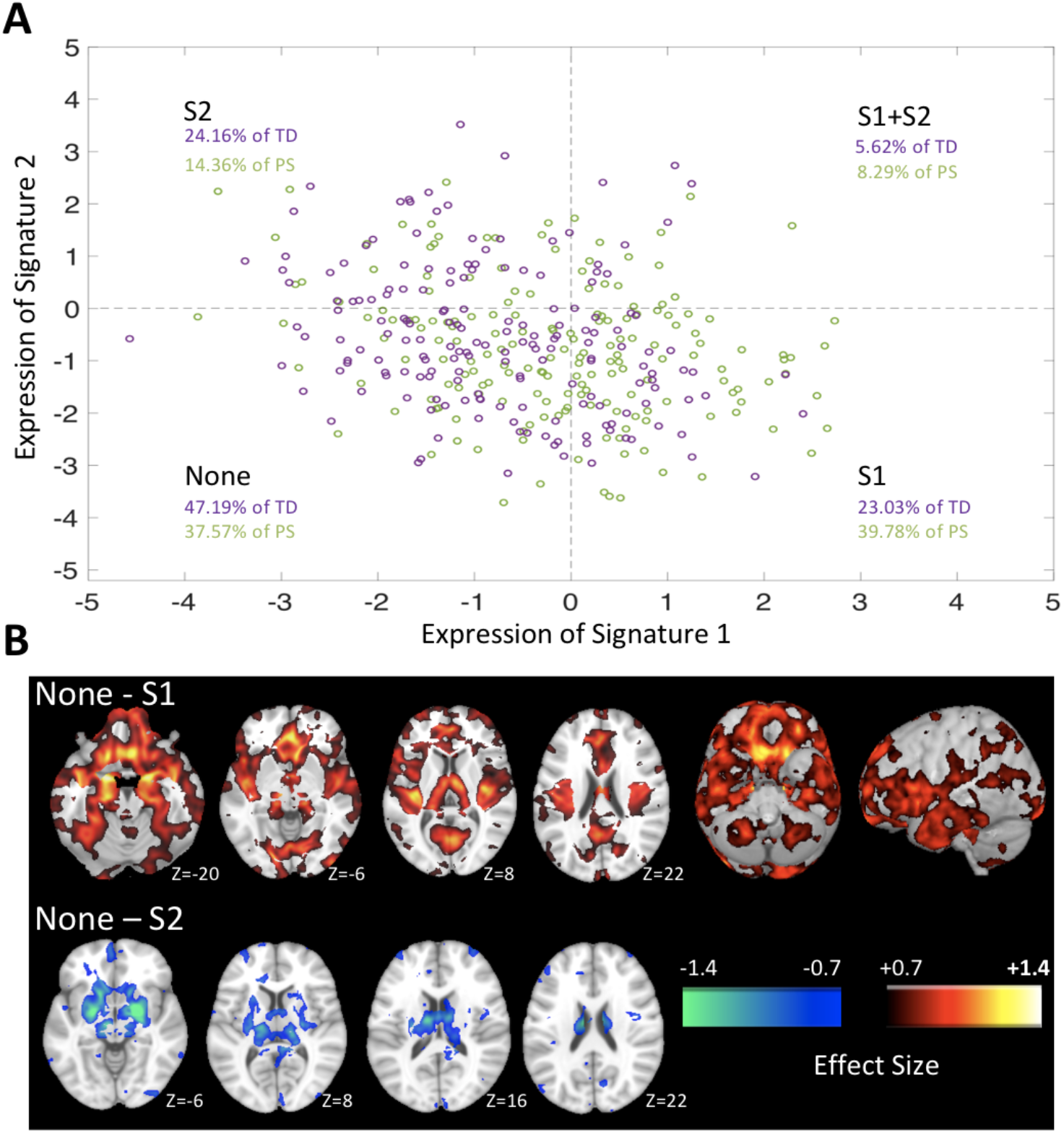
Presence of signature 1 (S1) and signature 2 (S2) expressions in the PNC dataset. **A**) Dimensional expression strength was visualized in a two-axis (E_1,_ E_2_) framework. TD and PS individuals across S1, S2, both signatures (S1+S2), and none of these two signatures (None). S1 was significantly more common in PS than in TD (*chi-square = 11*.*67; d*.*f. = 1; p < 0*.*05;* corrected for multiple comparisons) but other combinations were similar between PS and TD (p > 0.05). **B**) Voxel-based comparisons of regional gray matter volumes between individuals expressing primarily S1 (top) or S2 (bottom), compared to individuals expressing none of these two signatures, are displayed for visualization purposes. S1 individuals were characterized by significantly reduced gray matter volumes especially in prefrontal, temporal, and peri-Sylvian regions, whereas S2 individuals were characterized by markedly increased striatal volumes and normal-to-mildly enlarged cortical volumes compared to ‘None’ subgroup. Cohen’s d (effect size) maps were generated by masking MIDAS results after FDR-correction over voxels at p < *0*.*05*, and the largest effect sizes (∼1.4) were observed in the thalamus, nucleus accumbens, medial temporal, medial prefrontal/frontal and insular cortices for S1 and in the striatal region for S2. (See **Figure S3** for visualization of white matter comparison).

In PS, prevalence of signature 1, signature 2, both signatures, and ‘None’ was 39.78%, 14.36%, 8.29%, 37.57%, respectively. PS youths displayed significantly higher signature 1 prevalence than TD youths (*chi-square = 11*.*67; d*.*f. = 1; p < 0*.*05*), whereas the frequency difference between PS and TD was not significant for signature 2, both signatures, or ‘None’. Descriptive voxel-wise group comparisons were carried out between signature 1 or signature 2 and ‘None’ in order to visually illustrate the signature patterns. Signature 1 was characterized by widespread reductions in GM and WM volumes, while signature 2 was characterized by generally normal regional brain volumes except for markedly increased striatal and internal capsule volumes (**Figure 1B**; **Figures S3-S4**; Cohen’s d effect sizes ∼1.4 in regions showing most prominent differences). Within signature 1 and signature 2 groups, the pattern of GM volume did not differ significantly between PS or SZ and their respective control groups (**Figure S5**; see **Figure S6** for exploratory comparisons). However, the expression strength of signature 1 and signature 2 was higher for SZ than normal controls (NC), without a statistical difference between PS and TD (**Figure S7**). Exploratory analyses in a separate PNC subsample with other non-PS psychopathology are presented in **supplement** (“SZ signature prevalence in other psychopathology”). Exploratory analyses in TD and PS participants with age >=10 years and below 16 years are also presented (**supplement** “Schizophrenia signature prevalence in PNC younger participants”).

### Associations between signature 1, signature 2, cognition, and psychosis-spectrum symptoms in the PNC

Across the full PNC dataset (TD and PS) (**Figure 2 A-D**), signature 1 expression correlated inversely with cognitive efficiency (combination of accuracy and speed) (Spearman’s correlation; *ρ = -0*.*28; p < 10*^*-4*^), accuracy (*ρ = -0*.*24; p < 10*^*-4*^), and speed (*ρ = -0*.*16; p < 0*.*05)*, whereas stronger signature 2 expression correlated positively with efficiency (*ρ = 0*.*13; p < 0*.*05*) or speed (*ρ = 0*.*13; p < 0*.*05)* without significant accuracy correlation. Compared to PNC youths with signature 2, PNC youths with signature 1 had lower efficiency (Wilcoxon test; *z-stat = -4*.*64; p < 10*^*-4*^), accuracy (*z-stat = -3*.*38; p < 0*.*05*), and speed (*z-stat = -2*.*71; p < 0*.*05*) (**Figure 2 E-G**). Compared to those with neither SZ signature, signature 1 individuals also had lower efficiency (*z-stat = -4*.*50; p < 0*.*05*), accuracy (*z-stat = -3*.*91; p < 0*.*05*), and speed (*z-stat = -2*.*53; p < 0*.*05*). Consistent results were found after excluding individuals with threshold-level psychosis ratings (see **supplement** “SZ signatures excluding participants with subthreshold psychosis criteria”). Exploratory results for individual CNB tasks are presented in **Figure S8**. TD and PS were also explored separately, and similar signature-cognition relationships were found (**Figure S9)**. Exploratory results for available dimensional clinical measures are shown in **Tables S2-S3**. There was an inverse relationship between signature 1 and signature 2 expression strength (*ρ = -0*.*28; p < 10*^*-4*^).

**Figure 2.**
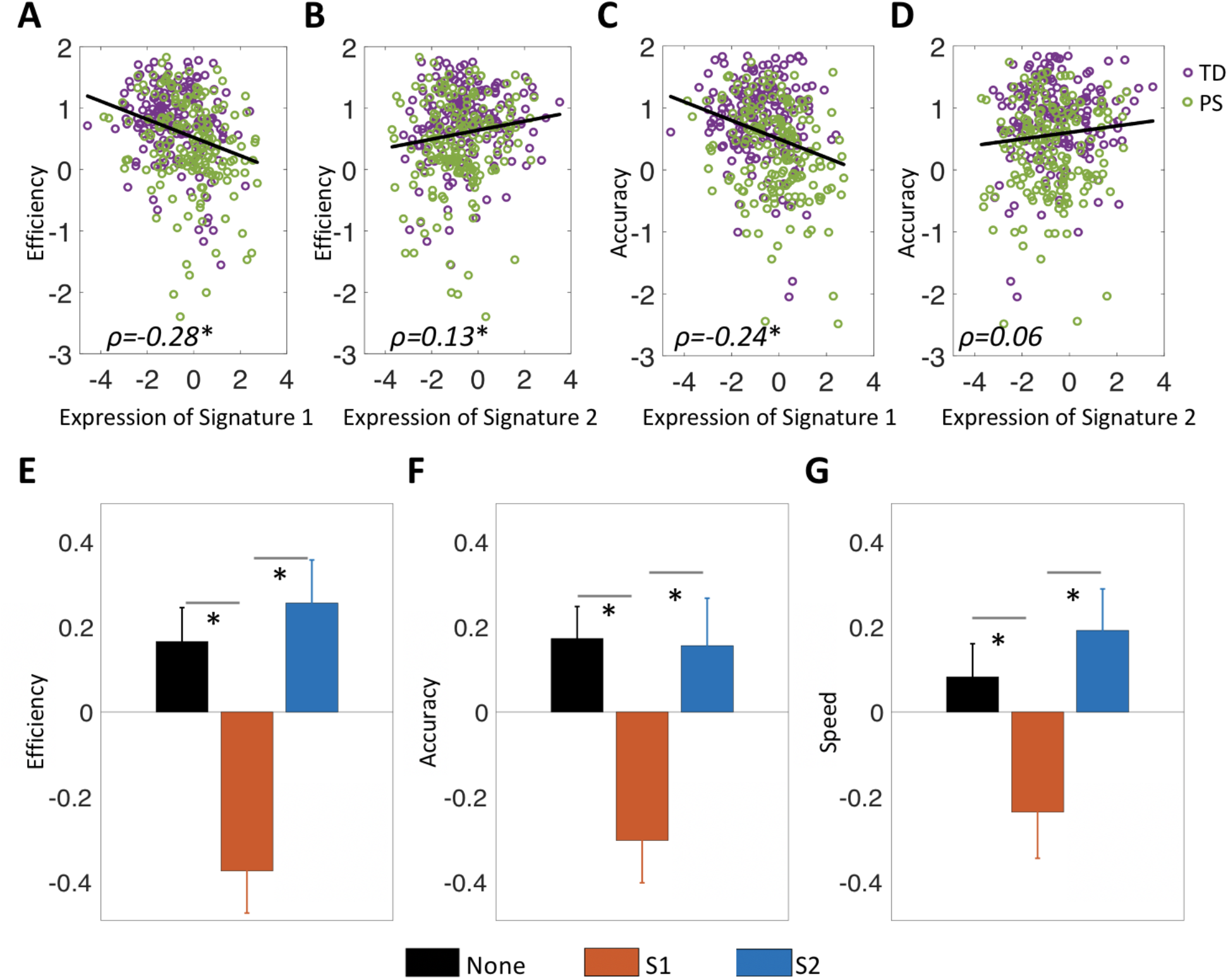
Cognitive profiles of signature 1 (S1) and signature 2 (S2) in the PNC dataset. **A-D**) S1 expression across the full sample correlated inversely with global cognitive performance efficiency (combination of accuracy and speed) (Spearman’s rank correlation; *ρ = -0*.*28; p < 10*^*-4*^) and accuracy (*ρ = -0*.*24; p < 10*^*-4*^), whereas stronger S2 expression correlated with higher efficiency (*ρ = 0*.*13; p < 0*.*05*) and S2 expression did not associate significantly with accuracy (*ρ = 0*.*06; p > 0*.*05*). **E-G**) S1 had worse efficiency (Wilcoxon rank sum test; *z-stat = -4*.*64; p < 10*^*-4*^), accuracy (*z-stat = -3*.*38; p < 0*.*05*), and speed (*z-stat = -2*.*71; p < 0*.*05*) than S2. S1 also had worse efficiency (*z-stat = -4*.*50; p < 0*.*05*), accuracy (*z-stat = -3*.*91; p < 0*.*05*), and speed (*z-stat = -2*.*53; p < 0*.*05*) than ‘None’. (Error bars represent Standard Error of Mean (SEM) over subjects; *: *p < 0*.*05;* corrected for multiple comparisons).

### Prevalence of SZ imaging signatures in the UKBB

In UKBB adults, prevalence of signature 1, signature 2, both signatures, and none of SZ signatures was 24.28%, 20.34%, 9.57%, and 45.81%, respectively (**Figure 3A**).

**Figure 3.**
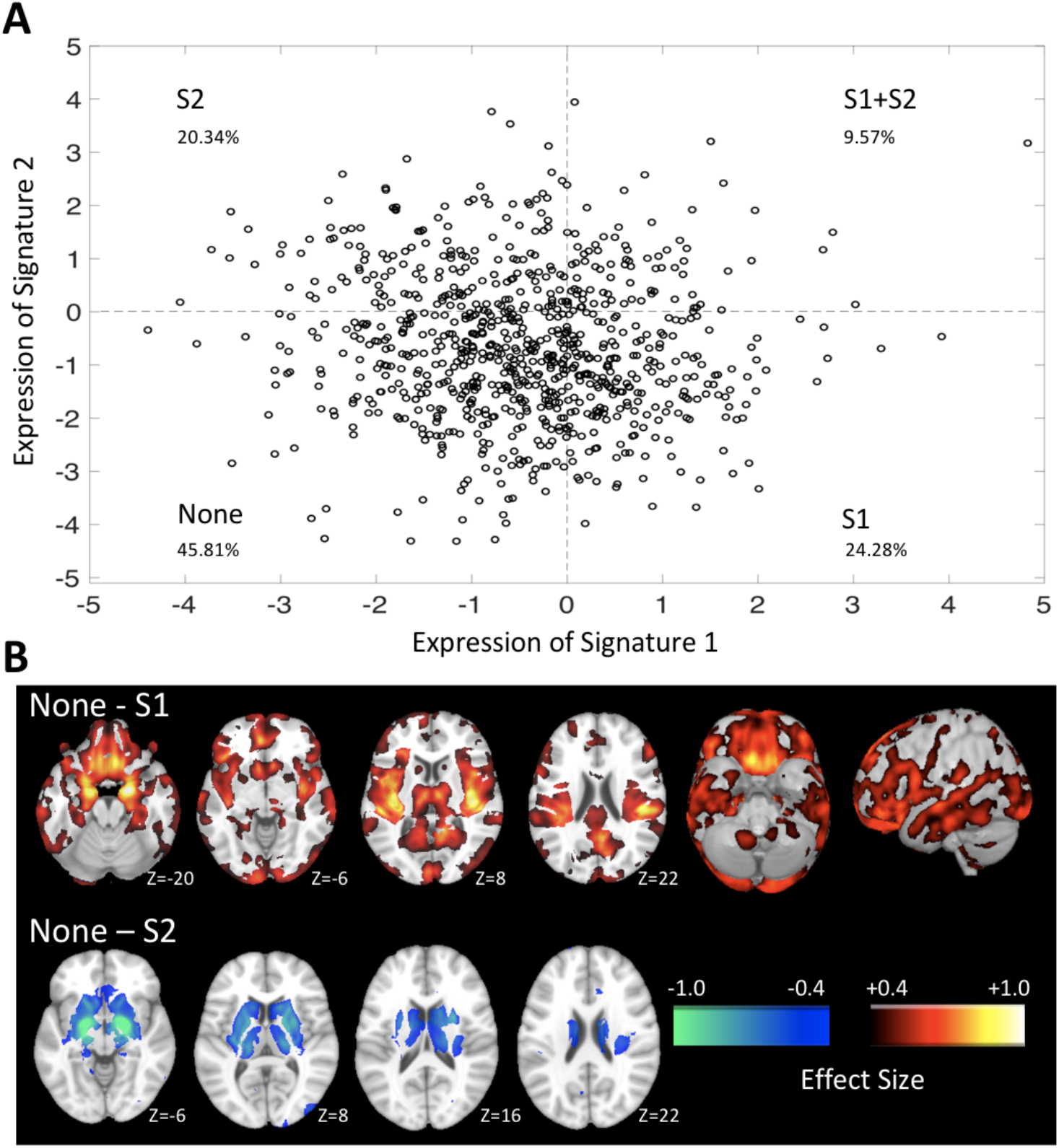
Presence of signature 1 (S1) and signature 2 (S2) expressions in the UKBB dataset. **A**) Dimensional expression strength was visualized in a two-axis (E_1,_ E_2_) framework. Individuals across S1, S2, both signatures (S1+S2), and none of these two signatures (None). **B**) Voxel-based comparisons of regional gray matter volumes between individuals expressing primarily S1 (top) or S2 (bottom), compared to individuals expressing none of these two signatures, are displayed for visualization purposes. S1 individuals were characterized by significantly reduced gray matter volumes especially in prefrontal, temporal, and peri-Sylvian regions, whereas S2 individuals were characterized by markedly increased striatal volumes compared to ‘None’ subgroup. Cohen’s d (effect size) maps were generated by masking MIDAS results after FDR-correction over voxels at p < 0.05. (See **Figure S10** for visualization of white matter comparison).

Signature 1 was characterized by widespread reductions in GM and WM volumes (compared to ‘None’), while signature 2 exhibited markedly increased striatal and internal capsule volumes (**Figure 3B**; **Figure S10**; Cohen’s d effect sizes ∼1.0 in regions showing most prominent differences). Exploratory analyses in the UKBB participants up to age 55 years are also presented in **supplement** (“Schizophrenia signature prevalence in UKBB older participants”).

### Associations between signature 1, signature 2, and cognition in the UKBB

In UKBB adults, greater signature 1 expression correlated with worse cognitive performance on the TMT-B task (*ρ = 0*.*15; p < 0*.*05*), with a non-significant correlation for signature 2 expression (**Figure 4A-B**). Signature 1 had worse TMT-B performance than signature 2 (*z-stat = 2*.*85; p < 0*.*05*) (**Figure 4C**; **Figure S11**). Fluid intelligence correlated negatively with signature 1 expression (*ρ = -0*.*15; p < 10*^*-4*^), but positively with signature 2 expression (*ρ = 0*.*17; p < 10*^*-4*^) (**Figure 4D-E)**, and fluid intelligence was lower in signature 1 than signature 2 (*z-stat = -5*.*16; p < 10*^*-4*^) and ‘None’ (*z-stat = -3*.*89; p < 0*.*05)*. Exploratory results for available dimensional clinical and other cognitive measures are shown in **Table S4** and **Figure S12**. Expressions of signature 1 and signature 2 were inversely associated (*ρ = -0*.*11; p < 0*.*05*).

**Figure 4.**
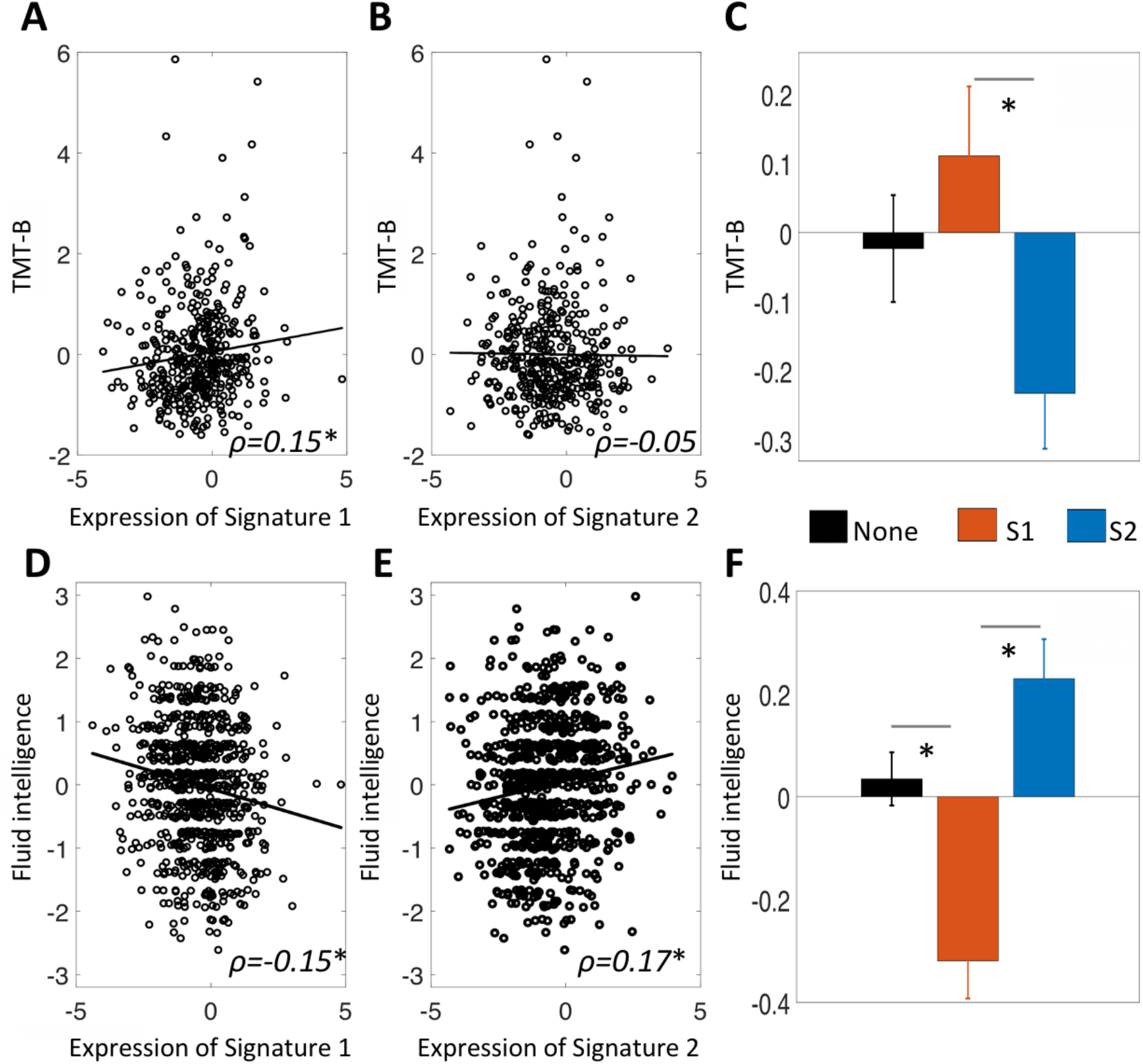
Cognitive profiles of signature 1 (S1) and signature 2 (S2) in the UKBB dataset. **A**) Stronger S1 expression across the full sample associated with worse cognitive performance on the trail making test part-B (TMT-B; note: higher TMT-B time reflects worse performance) (*ρ = 0*.*15; p < 0*.*05*), whereas **B)** S2 expression did not correlate significantly with TMT-B (*ρ = -0*.*05; p > 0*.*05*). **C**) S1 had worse cognitive performance on TMT-B than S2 (*z-stat = 2*.*85; p < 0*.*05*). **D**) Stronger S1 expression across the full sample correlated with worse cognitive performance on the fluid intelligence test (*ρ = - 0*.*15; p < 10*^*-4*^), whereas **E)** S2 expression correlated positively with the fluid intelligence test (*ρ = 0*.*17; p < 10*^*-4*^). **F**) S1 had worse cognitive performance on the fluid intelligence test than S2 (*z-stat = -5*.*16; p < 10*^*-4*^) and ‘None’ (*z-stat = -3*.*89; p < 0*.*05)*. (Error bars represent Standard Error of Mean (SEM) over subjects; *: *p < 0*.*05;* corrected for multiple comparisons).

### Genetic associations of signature 1 and signature 2 in the UKBB

In UKBB, signature 1 had significantly higher PRS_SZ_ compared to ‘None’ (Wilcoxon test; *z-stat = 2*.*69; p < 0*.*05*), but this was not seen for signature 2 (**Figure 5**; **Figure S13**; see **Figure S14** for dimensional associations between signature expression and PRS_SZ_).

**Figure 5.**
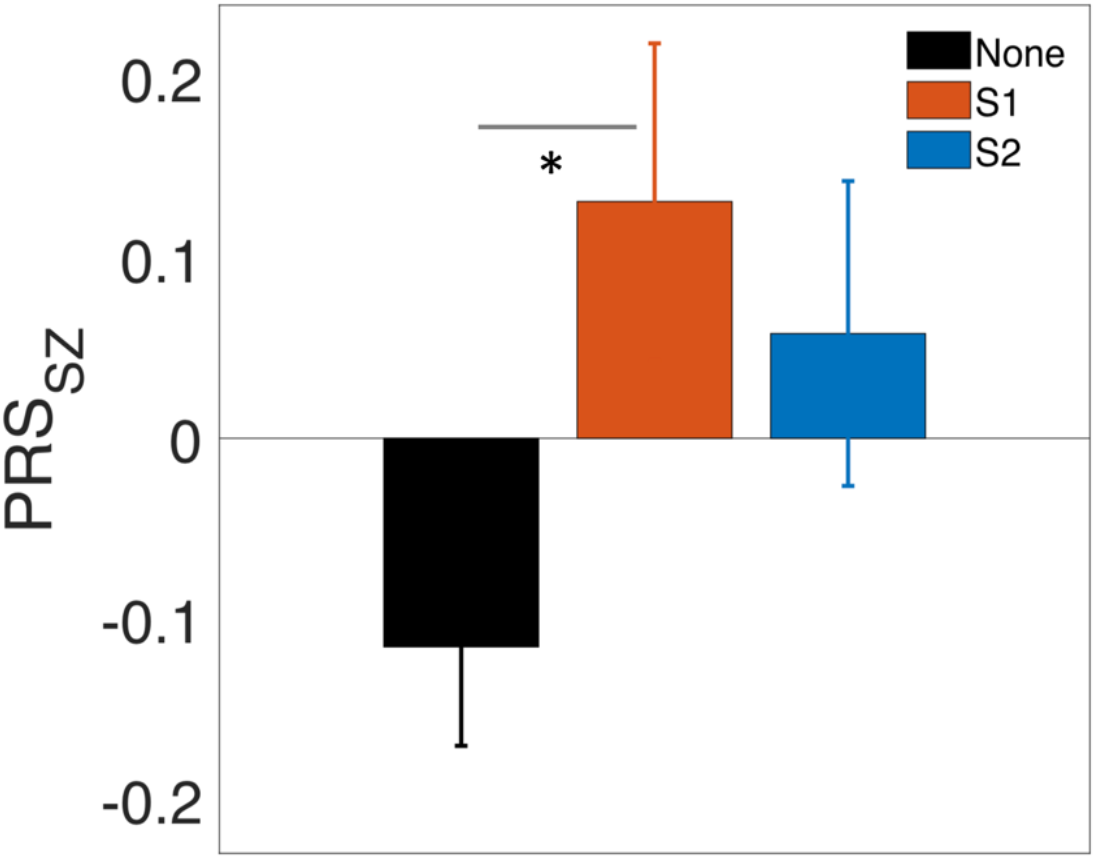
Genetic profiles of signature 1 (S1) and signature 2 (S2) in the UKBB dataset. Only S1 had significantly higher SZ polygenic risk scores (PRS_SZ_) compared to ‘None’ (Wilcoxon rank sum test; *z-stat = 2*.*69; p < 0*.*05*). (Error bars represent Standard Error of Mean (SEM) over subjects; *: *p < 0*.*05;* corrected for multiple comparisons).

## DISCUSSION

We replicated our recent findings of two novel neuroanatomical signatures of SZ (5), in an independent schizophrenia sample. We then examined the expression of these signatures in separate youth and adult population-based cohorts to measure signature prevalence, quantitative expression strength, and associations with psychosis-spectrum status, cognition, and genetics. Overall, moving from control to PS to SZ, the prevalence of both SZ signatures trends upward, consistent with these signatures reflecting psychosis-spectrum pathophysiology. Signature 1, marked by lower cortical GM and WM volumes, was associated with PS status in PNC youth and with impaired cognition in both PNC youth and UKBB adults. In contrast, the signature 2, marked by larger striatal and internal capsule volumes and normal-to-mildly-larger cortical GM and WM volumes, was not significantly associated with PS, and showed a positive association with cognitive performance. PRS_SZ_ were not significantly different between signature 1 and signature 2 individuals, and only signature 1 had significantly higher polygenic risk compared to individuals without either signature. A small fraction of healthy individuals (5-10%) expressed both signature 1 and signature 2, with somewhat higher percentages in PS than in TD youth. Thus, signature 1 and signature 2 capture distinct neurostructural dimensions, creating a novel two-axis framework for investigating the neuroanatomy of psychosis risk.

### Prevalence of SZ neuroanatomical signatures and their significance

As expected for dimensional brain imaging signatures, a substantial proportion of healthy individuals expressed them to some degree. The presence of SZ-related neuroanatomical signatures in healthy individuals raises the question of their clinical significance. It is possible that these signatures, particularly signature 1, might reflect neuroanatomical correlates of schizotaxia, a putative neurobiological state conferring risk for SZ (10, 35). Meehl, who developed the concept of schizotaxia, predicted that ∼10% of healthy individuals have schizotaxia. The idea that biological vulnerability to SZ is present in a substantial fraction of individuals even in the absence of a detectable pathological behavioral phenotype is highly plausible and supported by our findings here. Nonetheless, it is likely that only a minority of healthy participants identified here as expressing SZ-related signatures would have a substantially elevated risk of SZ. One possibility is that the signatures are truly dimensional, and that the lowest levels of expression are common but reflect negligible levels of pathophysiology/risk. Another possibility is that low levels of expression reflect normal variation in brain structure, which is etiologically unrelated to SZ, whereas at higher expression levels, perhaps above a specific threshold, its association with pathophysiology and vulnerability would be much stronger. Here we do see that the strength of signature 1 and signature 2 expression is larger in patients with SZ than in controls identified by the same binarized signature definitions. Yet another possibility is that healthy individuals expressing these signatures have other protective or compensatory structural or functional brain characteristics. Thus, the neuroanatomical signatures we identified in order to help parse the heterogeneity of SZ are themselves likely to have heterogeneity with respect to underlying etiological and microstructural contributors, coincidence of other biological and environmental factors, and hence pathophysiological significance. Even at a given level of expression, it is likely that the clinical significance of these signatures would depend on the population, for example varying by age and prevalence of various medical or psychiatric conditions, as these signatures are less robust at distinguishing PS from TD in youth than SZ from controls in adults. We note that the complexity of neuroanatomical signatures of risk is not substantially different from psychosis-risk approaches focused on subthreshold symptoms—population samples reveal such symptoms in 10-15% of youth, most of whom will not go on to develop a frank psychotic disorder (11, 36). While more work is required to clarify these issues, having a neurostructural approach to assess SZ risk complements the current symptom-based approach, and a combination of these (together with genetic and other biomarkers) is likely to optimize sensitivity and specificity for determining risk, establishing diagnosis, or predicting outcomes (37).

### Differences between signatures

Signature 1 was associated with PS status and with poorer cognition across groups, consistent with prior work linking reduced brain volume, especially in fronto-temporal regions, to worse cognitive performance, and with evidence of impaired cognition in SZ and at-risk states (17-19). Overall, signature 1 captures a more typical/prevalent SZ-related signature that is also more closely linked to known clinical risk factors in the general population. However, in PHENOM, signature 2 was present in about a third of individuals with established SZ and these individuals did not differ dramatically in clinical features from SZ with the signature 1 (aside from lower educational attainment in signature 1, and an association of signature strength with longer disease duration only in signature 1) (5). Signature 2 captures at least two aspects of brain structure—one is intact or mildly enlarged cortical volume, and the other is a marked enlargement of striatum and internal capsule. While it seems certain that a substantial subset of individuals with clinical risk for psychosis express a signature 2-like phenotype, the specificity of this signature 2 phenotype as defined here is not sufficient to identify a group with elevated psychosis-spectrum symptoms in the general population. Signature 1 may thus be more strongly associated with primary neurostructural abnormalities, while we speculate that clinical deficits in the signature 2 group, with its largely intact structure, are associated with primary functional abnormalities, perhaps in dopaminergic systems leading secondarily to basal ganglia enlargement. As noted above, a greater degree of signature 2 expression or combined occurrence with other factors may be needed to confer substantial SZ risk. Broadly, the results here as well as the prior findings in SZ are consistent with signature 1 reflecting an early-developmental vulnerability to SZ, while signature 2 reflects a signature devoid of substantial premorbid deficits.

The present study also identified increased polygenic risk for schizophrenia in individuals expressing signature 1 relative to those with neither signature; however polygenic risk did not differ between the two SZ signatures. These results are consistent with heterogeneity in the neurostructural phenotypes linked to the broad genetic risk profile captured by the PRS_SZ_. The more robust genetic relationship with signature 1 is consistent with higher prevalence of this signature in SZ; with prior evidence that polygenic risk is particularly associated with schizophrenia following early neurodevelopmental insults (22) and with cognitive impairment (23); and with findings in the general population linking SZ polygenic risk to lower cortical volume (14).

### Limitations

The present study has several important limitations which should be acknowledged. The data is cross-sectional, and future studies will need to assess change in signature expression over time, and whether signatures predict treatment response or other outcomes. The sample size for genetic analyses is relatively small; examination in larger samples will be important. While the PNC had a rich dataset of clinical and cognitive measures in youths and the UKBB had a rich dataset of genetics in adults, examination of large samples including comprehensive measures will be critical. Our clinical focus is on psychosis as these signatures were identified in schizophrenia, however a detailed understanding of the extent to which they are truly psychosis-specific will require future work across multiple psychiatric populations. Furthermore, this work only investigates neurostructural signatures; such volumetric measures are highly reliable, but an important future direction will be to examine noisier functional neuroimaging phenotypes (38), and evaluate multimodal biotypes. Lastly, we note that the relationships we identify have small effect sizes, as is typical for brain-behavioral relationships when accurately assessed in large samples (39); while this requires caution regarding expectations of clinical application, small effects can still provide critical clues regarding underlying pathophysiology (40).

## Conclusions

In summary, we identified two replicable SZ signatures, and show that they are also detectable in a majority of individuals with psychosis-spectrum symptoms even in the subclinical range, as well as a substantial minority of individuals without significant psychopathology. In these population-based cohorts, only the more neuroanatomically-abnormal signature 1 was associated with cognitive impairment, and the elevation in polygenic risk for SZ was somewhat higher in signature 1. These results may enhance future efforts to parse neurobiological heterogeneity and develop personalized approaches to identifying risk and preventing illness progression.

## Data Availability

All data in the present study are available upon reasonable request to the authors.

## Code availability

HYDRA (https://github.com/evarol/HYDRA) and MIDAS (https://github.com/evarol/MIDAS) codes used in this study are publicly available. The custom codes used in this study are also publicly available at https://github.com/ganchand/AJP_Codes.

