## Supplement for "Two schizophrenia imaging signatures and their associations with cognition, psychopathology, and genetics in the general population"

### Study samples

The PHENOM sample comprises individuals (age range: 16-45 years) with established schizophrenia (n = 307) and healthy controls (n = 364) from our previous study (1) that includes data from the USA, Germany, and China. USA participants were recruited at the University of Pennsylvania and provided written informed consent under a protocol approved by the Institutional Review Board. Participant assessment was conducted by expert clinical investigators. Diagnostic assessment employed the Structured Clinical Interview for DSM-IV (SCID) and included review of clinical records. Participant exclusion criteria were a history of substance abuse in the past six months or a positive urine drug screen on the day of the study and medical or neurological disorder that may impact brain function. Healthy control participants were excluded if they met criteria for any DSM-IV psychiatric disorder. Scale for the Assessment of Positive Symptoms (SAPS) (2) and the Scale for the Assessment of Negative Symptoms (SANS) (3) were assessed for patients. In Germany, participants were recruited at Ludwig-Maximilians University following study approval from the ethics committee. Participants provided written informed consent. Patient assessments were carried out by expert clinicians. Assessment included the SCID for Axis I & II disorders (SCID-I/-II), a semi-standardized clinical assessment of medical and psychiatric history, review of medical records and psychotropic medications, and the Positive and Negative Syndrome Scale (PANSS) (4) for disease severity and psychopathology. Individuals were excluded if they had other psychiatric and/or neurological diseases, past or present regular alcohol abuse, consumption of illicit drugs, past head trauma with loss of consciousness or electroconvulsive treatment, insufficient knowledge of German, IQ < 70, and age < 18 or > 65 years. Healthy controls with a positive familial history for mental illnesses (first degree relatives) were also excluded. In China, participants were recruited at Tianjin Medical University General Hospital following study approval from the ethics committee. Written informed consent was provided by each participant. Diagnosis of patients was assessed following the consensus of two clinical psychiatrists using DSM-IV/SCID. Inclusion criteria were 16–60 years of age and right-handedness. Exclusion criteria were MRI contraindications, pregnancy, history of systemic medical illness, central nervous system disorder and head trauma, and substance abuse within the last three months or lifetime history of substance abuse or dependence. For healthy control participants, the exclusion criteria were a history of psychiatric disease and first-degree relatives with a psychotic disorder. PANSS scores were assessed for patients for disease severity and psychopathology.

The PNC is a large community-based collaborative study between the University of Pennsylvania and the Children's Hospital of Philadelphia (5, 6). There were 9,498 participants who underwent clinical and cognitive phenotyping. Participants were proficient in English language and had no significant intellectual disability. Participant medical exclusion criteria were implanted metal, claustrophobia, malignancy, cystic fibrosis, immune disorders, renal or hepatic conditions, sickle cell anemia, epilepsy, stroke, head injury with loss of consciousness for more than five minutes, thyroid abnormalities, or other endocrine disorders. Medically healthy participants were selected for cross-sectional multimodal neuroimaging (n = 1,601, age 8-23 years) from the pool of clinically- and cognitively- assessed participants after stratifying for age and sex. Clinical assessment of participants was carried out using a comprehensive computerized tool for structured evaluation of psychopathology domains (GOASSESS) (6, 7). There were 399 participants who met psychosis spectrum symptoms (PS) criteria while 428 were typically developing (TD) individuals. As in Calkins et al. (8, 9), we identified participants as PS if they: 1) had an age-deviant PRIME (10) total score >= 2SD above age matched peers or had >= one PRIME item rated 6 or >= three items rated 5 (Somewhat Agree); 2) endorsed definite or possible hallucinations or delusions on the K-SADS psychosis screen (11); or 3) had an age-deviant total negative/disorganized SIPS (12) score >= 2SD above age-matched peers. TD participants lacked any significant psychopathology, and any history of psychoactive medication use or inpatient psychiatric treatment. After restricting the PNC sample to participants age >= 16 years old (to ensure based overlapping age with the PHENOM participants), our final PNC sample consisted of 181 PS and 178 TD individuals.

The UKBB is a large-scale study and an open access resource for neuroimaging, genetic, and cognitive datasets (13, 14). We selected UKBB participants from the 22,000 with neuroimaging released by 2019, with age below 50 years, based on their overlapping age with PHENOM. Among those n = 836, we used the ICD-9 codes (Field IDs 41271, 41203, 41205), ICD-10 codes (Field IDs 41270, 41202, 41204), and self-report (Field ID 20002) to determine the psychological disease burden on the study population. Included in our cohort, there are: zero participants diagnosed with schizophrenia (ICD-10 = {F20.0-F20.9}, ICD-9 = {2941, 2900-2909}, self-report = {1289}); zero participants diagnosed with autism (ICD-10 = {F84.0-F84.1}); zero participants diagnosed with Asperger's disease (ICD-10 = {F84.5}); zero participants diagnosed with dementia/Alzheimer's disease (ICD-10 = {F00.0-F00.9}, ICD-9 = {2950-2959}); fifty-four participants with self-reported depression (self-report = {1286}); twelve participants with self-reported anxiety (self-report = {1287}); one participant with self-reported bipolar disorder (self-report = {1291}); two participants with self-reported history of self-harm (self-report = {1290}); and two participants with self-reported history of nervous breakdown (self-report = {1288}). Medication status (whether participants were ever prescribed a medication for unusual or psychotic experience) was also studied (Field ID 20466.0.0). Fluid intelligence (Field ID 20016.2.0) (available in 94.62% samples) and TMT-B (Field ID 6350.2.0) (available in 46.53% samples) cognitive tests were included in the main analysis based on availability of sample size. We performed exploratory analysis for digit symbol substitution (Field ID 23324.2.0) (19.38% available), numeric memory (Field ID 4282.2.0) (22.37% available), and matrix pattern completion (Field ID 6373.2.0) (19.38% available) due to small sample size. European ancestry was determined in participants by the UKBB study team. The UKBB provided a binary descriptive field in the UKBB dataset designating the status of genetically inferred Caucasian (Field ID 22006.0.0). Related individuals were not included in the analyses. Identical by descent (IBD) estimation was done using a threshold of 0.25 to remove up to second degree relatives. There were n = 671 individuals with European ancestry; this subset was used in the genetic analysis. This research has been conducted using the UKBB Resource under Application Number 35148.

For replication of recently found SZ subtypes (1), we combined data from two previous study samples—Melbourne (Australia) (15) and Utrecht (Netherlands) (16). The local Research and

Ethics Committee in each location approved the study. In Melbourne data, there were 59 participants with SZ and 83 healthy volunteers. SZ diagnoses were based on SCID and chart review using DSM criteria (DSM-III-R, DSM-IV). Healthy volunteers were recruited through advertisements and they had similar socio-demographic areas as patients. All participants were screened for co-morbid medical and psychiatric conditions by clinical assessment, physical and neurological examination. Exclusion criteria were a history of significant head injury, seizures, electro-convulsive therapy during the six months prior to scan, polydipsia, neurological diseases, impaired thyroid function, steroid use or DSM criteria of alcohol or substance abuse or dependence. In Utrecht data, there were 90 participants with SZ and 115 healthy volunteers. SZ patients met DSM-IV criteria for schizophrenia and healthy controls met Research Diagnostic Criteria of never [being] mentally ill. The presence or absence of psychopathological abnormality was assessed by at least one independent trained rater. All participants provided written informed consent. The study was reviewed and approved by the local Medical Research and Ethics Committee of the University Medical Centre Utrecht.

It is worth noting that the PNC age ranges 8-23 years, but we restricted it to 16 years and above in the main analyses because it would be challenging to harmonize the data if age range does not overlap or is very different between data sites. To include a reasonable number of samples (n = 836, where 671 subjects were with European ancestry) from UKBB, we stretched the age limit up to 50 years. We did not study age beyond 50 years in the main analyses as this would impede our harmonization steps with PHENOM sample age ranges. In the replication study, where we did not need to harmonize data with PHENOM, we had age ranges from 16 to 57 years.

**Clinical measures**

To compare clinical profiles between signatures, GOASSESS (6, 7) psychopathology measures were studied in PNC. To provide a dimensional summary of the diverse psychopathology, a confirmatory factor analysis (17) was performed to orthogonally model four factors (mood, psychosis, externalizing, and phobias/fear) plus a general factor—overall psychopathology—that represents the symptoms across all psychiatric classifications. The mood, psychosis, externalizing, and phobias/fear specific dimensions were also examined without orthogonalization using a correlated factors model. As these models combined all psychotic symptoms into single factors, we also explored psychosis-specific models where psychosis symptoms were modelled with a single overall factor, with two factors (positive and negative), and with three factors (positive, positive-delusion, and negative).

**Image acquisition and preprocessing**

In site 1 of PHENOM (USA), structural images were acquired on a 3T TIM TRIO scanner (Siemens, Erlangen, Germany) using T1-weighted 3D magnetization-prepared rapid acquisition with gradient echo sequences (MPRAGE) using the TR = 1810 ms, TE = 3.51 ms, TI = 1100 ms, flip angle = 9 degree, FOV = 240 mm x 180 mm, matrix = 256 × 192, slices = 160, and slice/skip thickness = 1 mm/0 mm. In site 2 of PHENOM (Germany), T1-weighted MPRAGE images were acquired on a 1.5 T Magnetom Vision scanner (Siemens, Erlangen, Germany) using the TR = 11.6 ms, TE = 4.9 ms, FOV = 230 mm, matrix = 512 x 512, 126 contiguous axial slices of 1.5 mm thickness, and voxel size = 0.45 x 0.45 x 1.5 $mm^3$. In site 3 of PHENOM (China), images were acquired on a 3T MR system (Discovery MR750, General Electric, Milwaukee, WI, USA). Sagittal 3D T1-weighted images were acquired using a brain volume sequence (BRAVO) with the following parameters: TR = 8.2 ms, TE = 3.2 ms, TI = 450 ms, flip angle = 12 degree, FOV = 256 mm x 256 mm, matrix = 256 x 256, slice thickness = 1 mm, no gap, and 188 sagittal slices. In PNC, structural images were acquired on a Siemens 3T Tim Trio using MPRAGE with the following parameters: TR = 1810 ms, TE = 3.51 ms, TI = 1100 ms, FOV = 180 mm x 240 mm, matrix = 256 x 192, slices = 160, flip angle = 9 degrees, voxel size = 0.9 x 0.9 x 1$mm^3$. In UKBB, structural images were acquired on a Siemens 3T Skyra using MRPRAGE with the following parameters: TR = 2000 ms, TE = 2.01 ms, TI = 880 ms, FOV = 256 mm x 256 mm, slices = 208, flip angle = 8 degrees, voxel size = 1 x 1 x 1$mm^3$. In Melbourne (Australia) data, participants were scanned using the GE Signa 1.5 T scanner (GE, Milwaukee). A 3D volumetric spoiled-gradient recalled echo in the steady state sequence generated 124 continuous, 1.5 mm coronal slices. The following imaging parameters were used: TE = 3.3 mm, TR = 14.3 mm, flip angle =30 degree, matrix size = 256 x 256, FOV = 240 mm x 240 mm, voxel dimension = 0.94 x 0.94 x 1.5 $mm^3$. . In Utrecht (Netherlands) data, structural MRI scans of the whole brain were obtained on a 1.5 T Achieva scanner (Philips, Netherlands). 3D T1-weighted coronal spoiled-gradient echo scan of the whole head (256 × 256 matrix, TE = 4.6 ms, TR = 30 ms, flip angle = 30 degree, 160–180 contiguous slices; 1 × 1 × 1.2 $mm^3$ voxel size, FOV = 256 mm) was acquired. Across all sites, images were quality checked for motion, image artifacts, or restricted field-of-view, and corrected for magnetic field inhomogeneity (18). MUSE (19) was used to segment each individual's images into anatomical ROIs, which consist of GM, WM, and CSF regions, and ROI volumes were computed. Voxel-wise regional volumetric maps (20) for GM and WM were generated by registering skull-stripped T1-images to MNI common space using a deformable registration method (21).

**Inter-site image harmonization and correction of covariates**

Potential site differences between images could be driven by a variety of factors, including scanners and acquisition parameters. Indeed, the literature includes methods for image-level harmonization using advanced machine learning/deep learning models aiming to remove variations by the parameters through complex nonlinear transformation in the data distribution (22, 23). However, such models often require repeated images scanned under various scanner parameters and often do not protect for biological variations driven by different sample characteristics and demographics across sites. Such models would be challenging to apply with the number of site/scanner parameter combinations included in our data. For our analyses, we have rigorously preprocessed imaging data using the MUSE (19) pipeline, extracted ROI volumes, and spatially registered voxel-wise regional volumetric maps (20) using the same spatial resolution (MNI space) for all sites and all subjects. By design, this method utilizes an ensemble of atlases coming from different scanners, field strengths, and acquisition protocols, which renders the method quite robust to such confounds compared to other methods (24). Hence, instead of trying to correct for all combinations of scanner/protocol parameters, we chose to use regression-based statistical harmonization models and estimate site effects by accounting for site indicators. This is to: 1) deal with the remaining systematic error on the top of the processing steps involved in the MUSE and voxel-wise regional volumetric maps pipelines; 2) allow explicit estimates of site differences as well as other covariate effects. Some state-of-the-art approaches using statistical harmonization (rather than image-level parameters harmonization) are the ComBat-related methods (25, 26), which explicitly model the site effects as additive and multiplicative on the errors. We did not employ ComBat methods, because exploratory analysis indicates that our original PHENOM data did not show significant variance differences in the scales of the ROI volumes with PNC (Brown Forsythe's test, $p = 0.12$) and UKBB (Brown Forsythe's test, $p = 0.50$), but only in the mean shifts. Hence to be consistent with our earlier study by Chand et al. (1) so that the controls-SZ subtype models can be applied adequately to the test datasets (PNC and UKBB) to estimate SZ signature expressions, we chose to perform a linear correction that only accounts for the additive site effects, but not the multiplicative site effects. This approach was consistently applied at both ROI and voxel-level. A schematic of our harmonization approach is presented below:

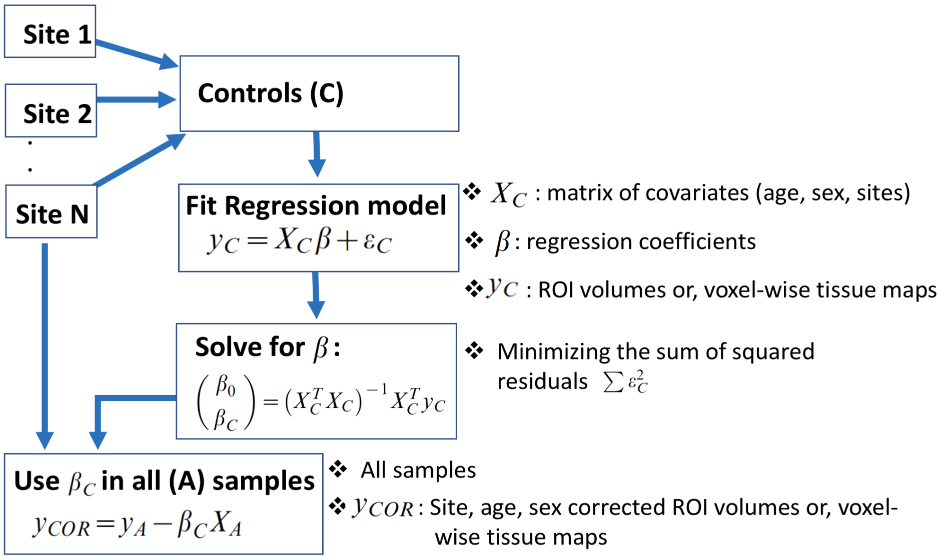

**Schematic of harmonization steps**: Regression coefficients for covariates (age, sex, sites) are computed from controls of all sites and then applied to data from each site to obtain the harmonized ROI volume or voxel-wise tissue (GM or WM) maps.

**Identifying the presence of SZ imaging signatures and defining subgroups**

Schizophrenia neuroanatomical dimensional model parameters—weight (W) and bias (b) parameters—were estimated from the PHENOM SZ and control samples using HYDRA (27). HYDRA parses disease heterogeneity by clustering cases through their associations with different faces of hyperplanes (decision boundaries) relative to the controls. HYDRA initializes the assignments of cases into clusters by sampling K-unit length hyperplanes considering the space of pairwise differences between cases and controls. The K-hyperplanes are sampled using determinantal point processes (28) and are used to estimate initial clustering assignments ($S^-$). While the estimated solution may vary depending on the initialization, a multi-initialization strategy is implemented. The final clustering results are accomplished based on a consensus of clustering solutions. The HYDRA algorithm can be summarized as:

**Input:** $X \in R^{n \times d}$, $Y \in \{-1, +1\}^n$ (training signals with 'n' participants, 'd' imaging features), K (number of subtypes)

**Output:** $W \in R^{d \times K}$, $b \in R^{1 \times K}$ (classifier); $S^-$ (clustering assignment)

**Initialization:** Initialize $S^-$

**Loop:** Repeat until convergence (or a fixed number of iterations)

Fix $S^-$, solve for W and b

Fix W and b, solve for $S^-$

The above analysis was carried out using 50 iterations between estimating hyperplanes and cluster estimation, 20 clustering consensus steps, and 0.25 regularization parameter. It is important to note that HYDRA internally performs many rounds of randomized K-fold leave-out procedures to assign subjects into clusters K-many times. Each of these K clustering results yields different sets of W's and b's that yield those clusters. HYDRA then performs a consensus step to harmonize the K different clustering assignments to generate a single cluster output for all subjects. In this step, the W's and b's that yielded the K clustering are lost. Therefore, to apply the HYDRA model to out of sample subjects, it is necessary to re-train the HYDRA decision boundary with the final clustering outputs to generate a final W and b to predict new samples. This last step does not change the optimal clusters in the training set but enables us to transfer this model to out of sample test data. HYDRA was applied to the PHENOM control and SZ samples using the volumes of 145 anatomical ROIs (**Table S1**), 10-fold cross-validation approach was performed to protect against overfitting, and the two optimal and distinct SZ dimensions were identified. The optimal parameters—"W" that consists of the weight of all ROIs and "b" that captures the associated bias term—that best separate the controls from the two SZ signatures were then applied to the harmonized PNC and UKBB datasets, respectively, where the same 145 ROIs volume features were considered. The test data (PNC or UKBB) were not touched during HYDRA training so that this procedure protects against overfitting. Mathematically, "expression" (E) of SZ signatures can be written via distance from respective hyperplanes of support-vector machine learning methods (27, 29) as,

$$E_i = W_i^T X + b_i$$

where, X is the feature matrix (ROI volumes) of PNC or UKBB participants. Since controls are assigned as '-1' and SZ patients as '+1' during HYDRA training, $E_i < 0$ indicates none of SZ signatures along dimension 'i', and $E_i > 0$ indicates SZ signature along dimension 'i'. Based on these expressions, each participant was thus assigned to one of these subgroups, S1 ($E_1 > 0$, $E_2 < 0$), S2 ($E_1 < 0$, $E_2 > 0$), None ($E_1 < 0$, $E_2 < 0$) and S1+S2 ($E_1 > 0$, $E_2 > 0$), and placed in a two-axis framework for visualization. Note, a participant in 'S1+S2' group has positive signatures along both dimensions ($E_1 > 0$, $E_2 > 0$). Typically, as in our original SZ analysis (1) or clustering studies (27) each participant is assigned to a single category based on the relative distance from hyperplanes. In the present study, however, in order to fairly compare proportions of samples across three studies (PNC, UKBB, and PHENOM), SZ signatures were estimated for PHENOM samples in the same way as in PNC and UKBB. Specifically, to evaluate the strength of SZ expressions, here we did not force the "mixed" participants into a single category but instead left them as 'S1+S2' participants, as we were interested in examining "pure" S1 or S2 subgroups by excluding the mixed subgroup. Although the 'S1+S2' subgroup is interesting, due to the small sample size of this group, especially in PNC dataset, the brain voxel-wise, cognitive, clinical, and genetic comparisons were mainly focused on 'None', 'S1' and 'S2' subgroups.

**MIDAS**

The voxel-wise volumetric profiles between the groups were assessed using MIDAS (30) for visualization purposes of signatures. MIDAS determines the optimal regional smoothing and provides higher sensitivity and specificity in identifying group alterations compared to other existing methods, such as standard voxel-based morphometry analysis using general linear models, where use of an *a priori* fixed smoothing kernel [e.g., 6-12 mm full width half maximum (FWHM) in general] is a prerequisite. To alert the reader that we are not using the more common (but we believe less-optimal) approach, we referred to our methodology as voxel-wise volumetric analysis instead of standard voxel-based morphometry. MIDAS parses any set of image maps using a sufficiently large set of overlapping neighborhoods (P), and performs regional discriminative analysis based on least squares support vector machines, which aims to relate the imaging features $X \in R^{n \times m}$ ('n' participants and 'm' dimensional imaging features) with group variable $Y \in R^n$ via a weight vector (w). The regional volumetric pattern that optimally discriminates between groups is equivalent to filtering locally by an optimal kernel whose coefficients are the weights of the discriminant. For a particular voxel 'i', statistic (S) is computed by modulating the total contribution of each voxel to the estimated local activation patterns (a) with the total predictive power of the respective machine learners:

$$s_i = \frac{\sum_{p=1}^{P} a_i^p}{\sum_{p=1}^{P} \| w^p \|_2^2}$$

where, $a \propto \frac{1}{n}(X - \bar{X})^T(X - \bar{X})w$, and $\sum_{p=1}^{P} \| w^p \|_2^2$ is the sum of the inverse predictive power of all learners, in which voxel 'i' participates. The statistical significance p-value corresponding to the voxel statistic is analytically achieved by approximating permutation tests. The voxel-wise regional volumetric maps (20) of GM or WM with dimensions 182 x 218 x 182 were used to evaluate the neuroanatomical differences between the groups. In MIDAS analysis, 15 neighborhood radii in voxels, 500 neighborhoods, and 0.1 regularization parameter were used. The voxel-wise statistical significance values (p-values) between signatures (subgroups) were extracted from MIDAS calculations, and false discovery rate (*FDR*) was used to correct for multiple comparisons across the voxels. The map of statistically significant regions ($FDR\text{-}p < 0.05$) was then used as a mask for the effect size (Cohen's d) maps in order to display the significant differences between signatures.

**SZ signatures excluding participants with subthreshold psychosis criteria**

We conducted additional sensitivity analyses, excluding the individuals rated as threshold severity on one or more GOASSESS psychosis items (n = 47 PS participants). In the remaining 134 PS, the patterns of results were very similar in terms of the pattern of effect sizes, although statistical significance was somewhat reduced as expected given lower sample size. In this reduced PS sample, the prevalence of signature 1, signature 2, both signatures, and 'None' was 38.81%, 17.16%, 8.21%, and 35.82%, respectively (in the full PS sample it was 39.78%, 14.36%, 8.29%, and 37.57%; see **Figure 1A** in main text). PS youths in the reduced sample still displayed significantly higher signature 1 prevalence than TD youths ($p < 0.05$), and the frequency difference between PS and TD was still not significant for signature 2, both signatures, and 'None'. The effect size of the relationship between CNB cognitive performance and the two signatures was also similar in the reduced PS sample (efficiency S1 $\rho = -0.22$, S2 $\rho = 0.15$; accuracy S1 $\rho = -0.21$, S2 $\rho = 0.05$; speed S1 $\rho = -0.14$, S2 $\rho = 0.17$) compared to the original PS sample (efficiency S1 $\rho = -0.25$, S2 $\rho = 0.12$; accuracy S1 $\rho = -0.25$, S2 $\rho = 0.02$; speed S1 $\rho = -0.14$, S2 $\rho = 0.15$). Moreover, across the new combined (TD+ reduced PS) sample, signature 1 expression remained negatively associated with global cognitive performance efficiency ($\rho = -0.25$; $p < 0.05$), accuracy ($\rho = -0.20$; $p < 0.05$), and speed ($\rho = -0.16$; $p < 0.05$), whereas stronger signature 2 expression correlated positively with higher efficiency (*$\rho = 0.13$; $p < 0.05$*) and speed (*$\rho = 0.13$; $p < 0.05$*) while positive correlation did not reach significance level with accuracy. Compared to signature 2, signature 1 had lower efficiency (*z-stat = -3.97; $p < 0.05$*), accuracy (*z-stat = -2.77; $p < 0.05$*), and speed (*z-stat = -2.44; $p < 0.05$*). Compared to those with neither SZ signature, signature 1 individuals also had lower efficiency (*z-stat = -4.20; $p < 0.05$*), accuracy (*z-stat = -3.48; $p < 0.05$*), and speed (*z-stat = -2.76; $p < 0.05$*). Signature 2 did not differ significantly from individuals with neither of SZ signatures in efficiency, accuracy, and speed. These results were consistent with the findings shown in **Figure 2** (main text). Thus, it is unlikely that our results are explainable by the minority of PS participants who may have frank psychosis.

**SZ signature prevalence in other psychopathology**

We explored other psychopathology individuals (n = 239, age 16-23 years) from the PNC who were neither TD nor PS (11). Prevalence of SZ signature 1, signature 2, both signatures, and none of SZ signatures ('None') was 27.62%, 22.18%, 9.2%, and 41%, respectively. Signature 1 prevalence in other psychopathology individuals (27.62%) was significantly lower than that of PS (39.78%) ($p < 0.05$), but slightly greater than that of TD (23.03%) with no significant difference. Signature 2 prevalence in other psychopathology individuals (22.18%) was not statistically different than that of TD (24.16%) or PS (14.36%) ($p > 0.05$). These findings provide some support for relative specificity for signature 1 since its prevalence is greater in PS than in those with "other psychopathology". Detailed studies will be needed to fully evaluate diagnostic and dimensional specificity in the future.

**Schizophrenia signature prevalence in PNC younger participants**

We further explored neuroanatomical schizophrenia signatures in PNC (TD and PS) participants below 16 years old. There were n = 393 participants >=10 years old and below 16 years old (185 typically developing and 208 psychosis-spectrum) in PNC. In TD individuals, prevalence of schizophrenia signature 1, signature 2, both signatures, and none of schizophrenia signatures ('None') was 22.16%, 23.24%, 5.41%, and 49.19%, respectively. In PS individuals, prevalence of signature 1, signature 2, both signatures, and 'None' was 39.42%, 16.83%, 9.62%, 34.13%, respectively. These results were consistent with the prevalence of schizophrenia signatures presented in **Figure 1A** for age 16-23 years. Although similar prevalence in both age ranges, these results should be interpreted with caution because the original PHENOM models were not derived from participants of this age group.

**Schizophrenia signature prevalence in UKBB older participants**

We also explored the neuroanatomical schizophrenia signatures in UKBB participants up to age 55 years (n = 3,844). Prevalence of schizophrenia signature 1, signature 2, both signatures, and none of schizophrenia signatures ('None') was 24.92%, 20.66%, 9.16%, and 45.26%, respectively, and these results were similar with the main results in **Figure 3A** (below age 50 years). Although results stayed similar with varied ages, these results should be interpreted with caution, mainly because original PHENOM "schizophrenia signatures" were not derived from individuals of this age range.

**Table S1:** MUSE (19) anatomical regions of interest (ROIs) used to derive SZ signature expressions (L: Left hemisphere; R: Right hemisphere; WM: White matter).

| | | |
|---|---|---|
| 3rd ventricle | Ventral diencephalon (R) | Anterior insula (L) |
| 4th ventricle | Ventral diencephalon (L) | Anterior orbital gyrus (R) |
| Accumbens area (R) | Cerebellar vermal lobules I-V | Anterior orbital gyrus (L) |
| Accumbens area (L) | Cerebellar vermal lobules VI-VII | Angular gyrus (R) |
| Amygdala (R) | Cerebellar vermal lobules VIII-X | Angular gyrus (L) |
| Amygdala (L) | Basal forebrain (R) | Calcarine cortex (R) |
| Brain Stem | Basal forebrain (L) | Calcarine cortex (L) |
| Caudate (R) | Frontal lobe WM (R) | Central operculum (R) |
| Caudate (L) | Frontal lobe WM (L) | Central operculum (L) |
| Cerebellum exterior (R) | Occipital lobe WM (R) | Cuneus (R) |
| Cerebellum exterior (L) | Occipital lobe WM (R) | Cuneus (L) |
| Cerebellum WM (R) | Parietal lobe WM (R) | Entorhinal area (R) |
| Cerebellum WM (L) | Parietal lobe WM (L) | Entorhinal area (L) |
| Hippocampus (R) | Temporal lobe WM (R) | Frontal operculum (R) |
| Hippocampus (L) | Temporal lobe WM (L) | Frontal operculum (L) |
| Inferior lateral ventricle (R) | Fornix (R) | Frontal pole (R) |
| Inferior lateral ventricle (L) | Fornix (L) | Frontal pole (L) |
| Lateral ventricle (R) | Anterior limb of internal capsule (R) | Fusiform gyrus (R) |
| Lateral ventricle (L) | Anterior limb of internal capsule (L) | Fusiform gyrus (L) |
| Pallidum (R) | Posterior limb of internal capsule including cerebral peduncle (R) | Gyrus rectus (R) |
| Pallidum (L) | Posterior limb of internal capsule including cerebral peduncle (L) | Gyrus rectus (L) |
| Putamen (R) | Corpus callosum | Inferior occipital gyrus (R) |
| Putamen (L) | Anterior cingulate gyrus (R) | Inferior occipital gyrus (L) |
| Thalamus proper (R) | Anterior cingulate gyrus (L) | Inferior temporal gyrus (R) |
| Thalamus proper (L) | Anterior insula (R) | Inferior temporal gyrus (L) |
| Lingual gyrus (R) | Occipital fusiform gyrus (L) | Subcallosal area (R) |
| Lingual gyrus (L) | Opercular part of inferior frontal gyrus (R) | Subcallosal area (L) |
| Lateral orbital gyrus (R) | Opercular part of inferior frontal gyrus (L) | Superior frontal gyrus (R) |
| Lateral orbital gyrus (L) | Orbital part of inferior frontal gyrus (R) | Superior frontal gyrus (L) |
| Middle cingulate gyrus (R) | Orbital part of inferior frontal gyrus (L) | Supplementary motor cortex (R) |
| Middle cingulate gyrus (L) | Posterior cingulate gyrus (R) | Supplementary motor cortex (L) |
| Medial frontal cortex (R) | Posterior cingulate gyrus (L) | Supramarginal gyrus (R) |
| Medial frontal cortex (L) | Precuneus (R) | Supramarginal gyrus (L) |
| Middle frontal gyrus (R) | Precuneus (L) | Superior occipital gyrus (R) |
| Middle frontal gyrus (L) | Parahippocampal gyrus (R) | Superior occipital gyrus (L) |
| Middle occipital gyrus (R) | Parahippocampal gyrus (L) | Superior parietal lobule (R) |
| Middle occipital gyrus (L) | Posterior insula (R) | Superior parietal lobule (L) |
| Medial orbital gyrus (R) | Posterior insula (L) | Superior temporal gyrus (R) |
| Medial orbital gyrus (L) | Parietal operculum (R) | Superior temporal gyrus (L) |
| Postcentral gyrus medial segment (R) | Parietal operculum (L) | Temporal pole (R) |
| Postcentral gyrus medial segment (L) | Postcentral gyrus (R) | Temporal pole (L) |
| Precentral gyrus medial segment (R) | Postcentral gyrus (L) | Triangular part of the inferior frontal gyrus (R) |
| Precentral gyrus medial segment (L) | Posterior orbital gyrus (R) | Triangular part of the inferior frontal gyrus (L) |
| Superior frontal gyrus medial segment (R) | Posterior orbital gyrus (L) | Transverse temporal gyrus (R) |
| Superior frontal gyrus medial segment (L) | Planum polare (R) | Transverse temporal gyrus (L) |
| Middle temporal gyrus (R) | Planum polare (L) | |
| Middle temporal gyrus (L) | Precentral gyrus (R) | |
| Occipital pole (R) | Precentral gyrus (L) | |
| Occipital pole (L) | Planum temporale (R) | |
| Occipital fusiform gyrus (R) | Planum temporale (L) | |

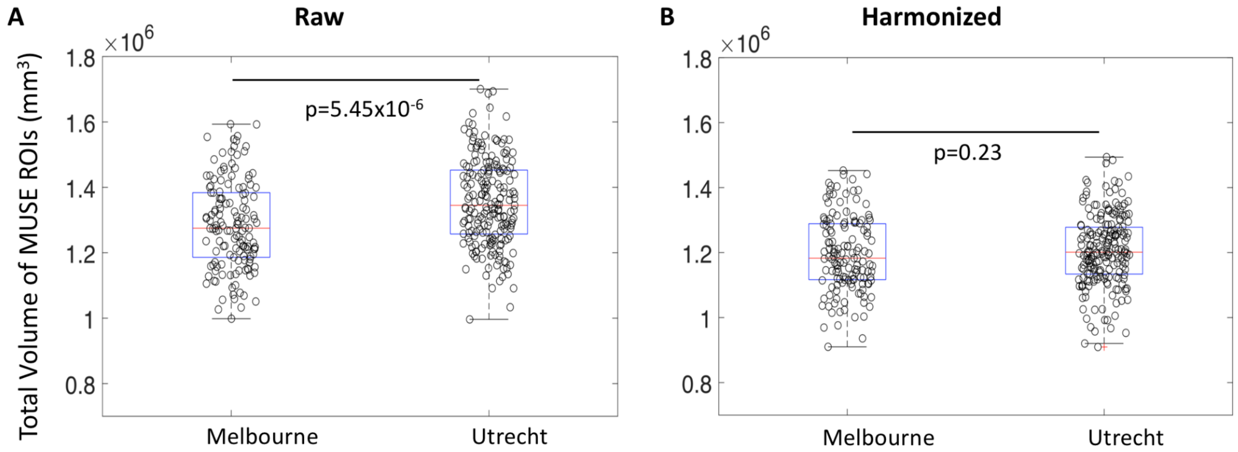


**Figure S1**: Validation datasets (n = 347; 198 healthy controls and 149 SZ patients) from Melbourne (Australia) and Utrecht (Netherlands). **A)** Total volume of MUSE ROIs before harmonization (Raw) where sites are significantly different ($p = 5.45 \times 10^{-6}$), and **B)** Total volume of MUSE ROIs after site, age, and sex correction (Harmonized) demonstrating our harmonization approach diminishes site differences ($p = 0.23$).

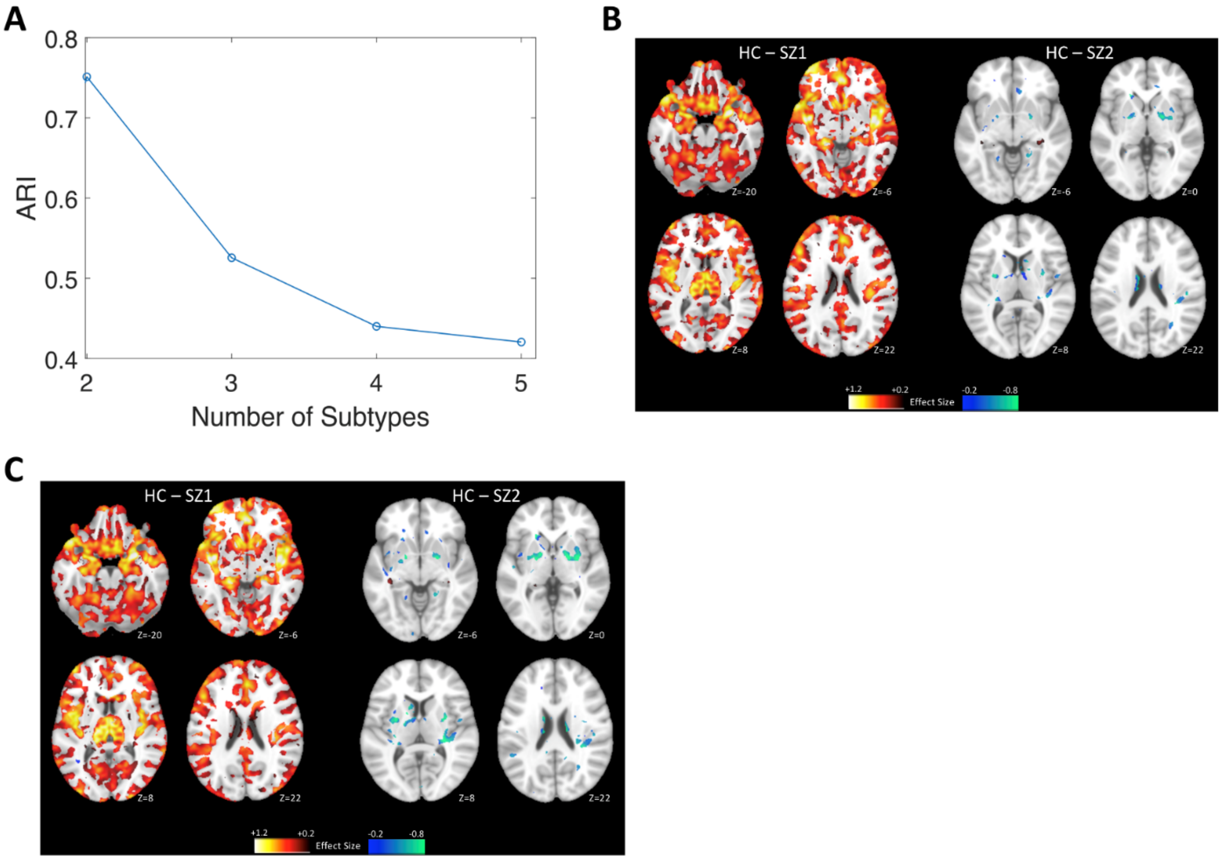


**Figure S2**: **Replication dataset. A)** HYDRA results demonstrating two SZ subtypes are highly stable [demonstrating a high adjusted Rand index (ARI) of 0.75] in independent datasets from Melbourne (Australia) and Utrecht (Netherlands), and **B)** MIDAS results demonstrate voxel-based comparisons of regional gray matter volumes between schizophrenia type 1 (SZ1) or type 2 (SZ2), compared to healthy controls (HC). Compared to HC, SZ1 individuals were characterized by widespread reduction of gray matter volumes, especially in the thalamus, nucleus accumbens, medial temporal, medial prefrontal/frontal and insular cortices, whereas SZ2 individuals were characterized by increased striatal volumes. These findings replicated our previous findings (1) in the present two independent datasets. **C) Reproducibility analysis**: Our previous models (1) were also applied to the replication data, and SZ1 and SZ2 were predicted. Voxel-based comparisons of regional gray matter volumes between HC and predicted SZ1 or SZ2 were consistent with B). (Cohen's d effect size maps were generated by masking MIDAS results after FDR-correction over voxels at $p < 0.05$).

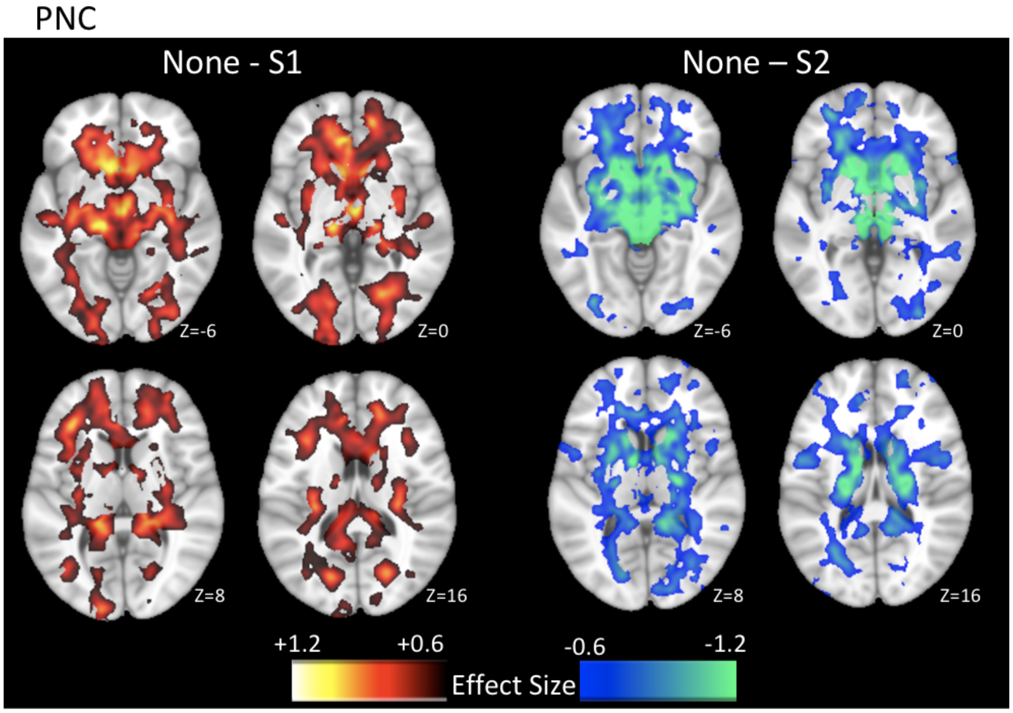


**Figure S3:** Voxel-based comparisons of regional white matter (WM) volumes between individuals expressing primarily signature 1 (S1) or signature 2 (S2), compared to individuals expressing none of these two signatures, in PNC youth are displayed for purely visualization purposes. S1 was characterized by widespread reductions of WM volumes, whereas S2 was characterized by increased WM volumes mainly in the internal capsule. (Cohen's d effect size maps were generated by masking MIDAS results after FDR-correction over voxels at $p < 0.05$).

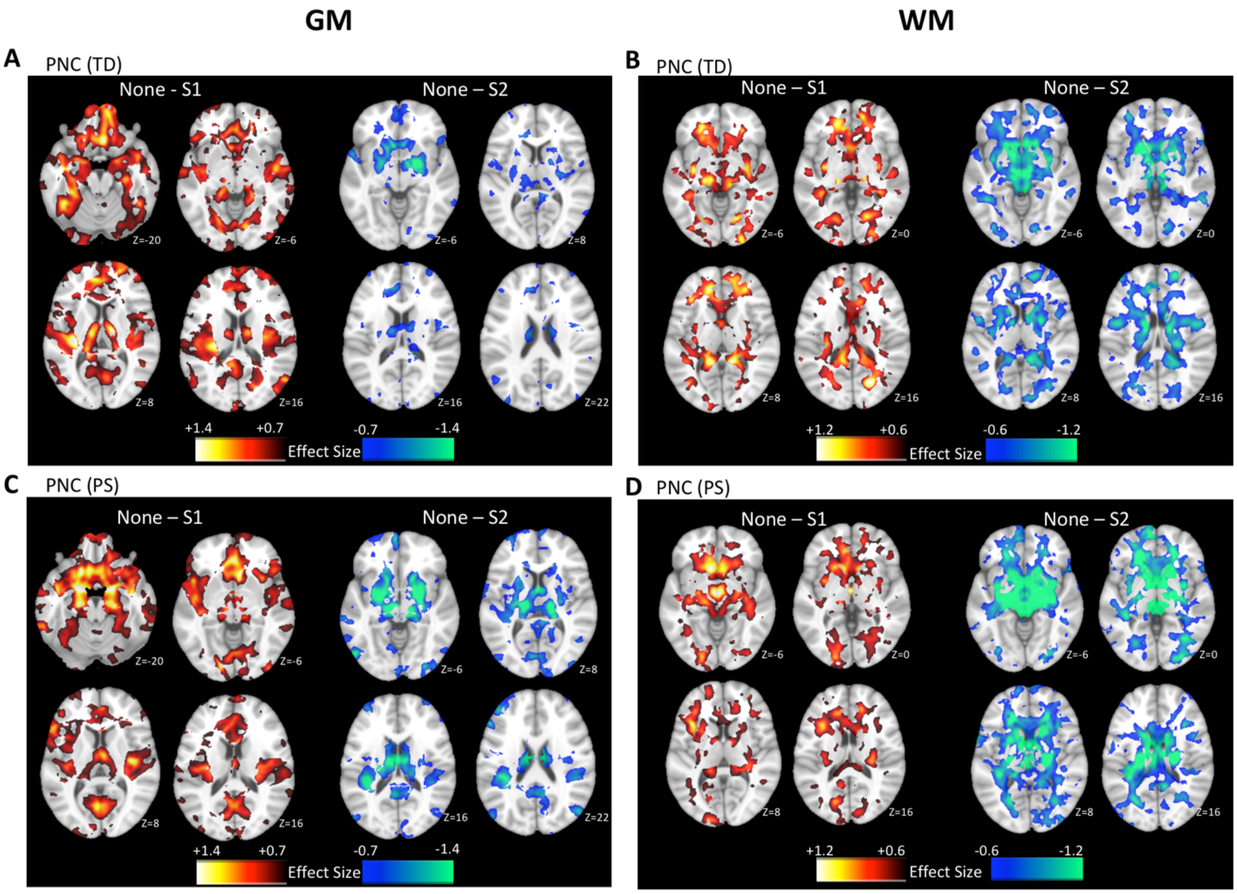


**Figure S4:** Voxel-based comparisons of regional gray matter (GM) or white matter (WM) volumes between individuals expressing primarily signature 1 (S1) or signature 2 (S2), compared to individuals expressing none of these two signatures, for TD and PS separately, are displayed for visualization purposes. S1 was characterized by significantly reduced GM volumes in prefrontal, temporal, and peri-Sylvian regions, whereas S2 was characterized by increased striatal volumes. S1 was also characterized by widespread reductions in WM volumes, whereas S2 was characterized by increased WM volumes primarily in the internal capsule. (Cohen's d effect size maps were generated by masking MIDAS results after FDR-correction over voxels at $p < 0.05$).

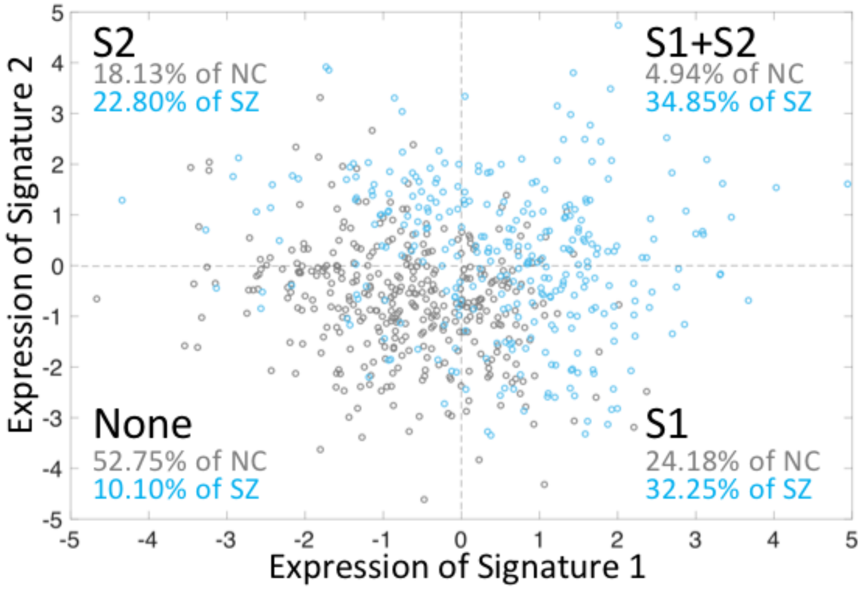


**Figure S5.** Prevalence of signature 1 (S1) and signature 2 (S2) expressions in the PHENOM samples consisting of SZ and normal controls (NC).

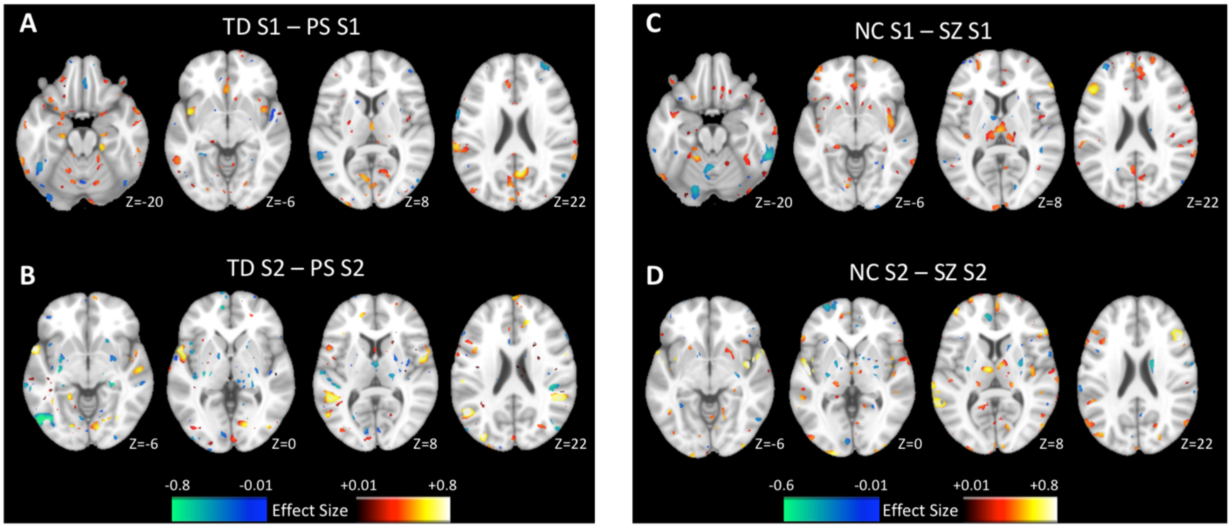


**Figure S6:** Voxel-based comparisons of regional GM volumes exploring potential protective/compensatory structural features in healthy individuals who express SZ-related signatures but do not exhibit psychotic symptoms. In PNC, **A)** Compared to PS S1, TD S1 was characterized by larger GM volumes in prefrontal, temporal, and peri-Sylvian regions, and **B)** Compared to PS S2, TD S2 was characterized by smaller striatal volumes but larger GM volumes in other regions. In PHENOM, **C)** Compared to SZ S1, NC S1 was characterized by larger GM volumes in prefrontal, temporal, and peri-Sylvian regions, and **D)** Compared to SZ S2, NC S2 was characterized by smaller striatal volumes but larger GM volumes in other regions. (Cohen's d effect size maps were generated by masking MIDAS results).

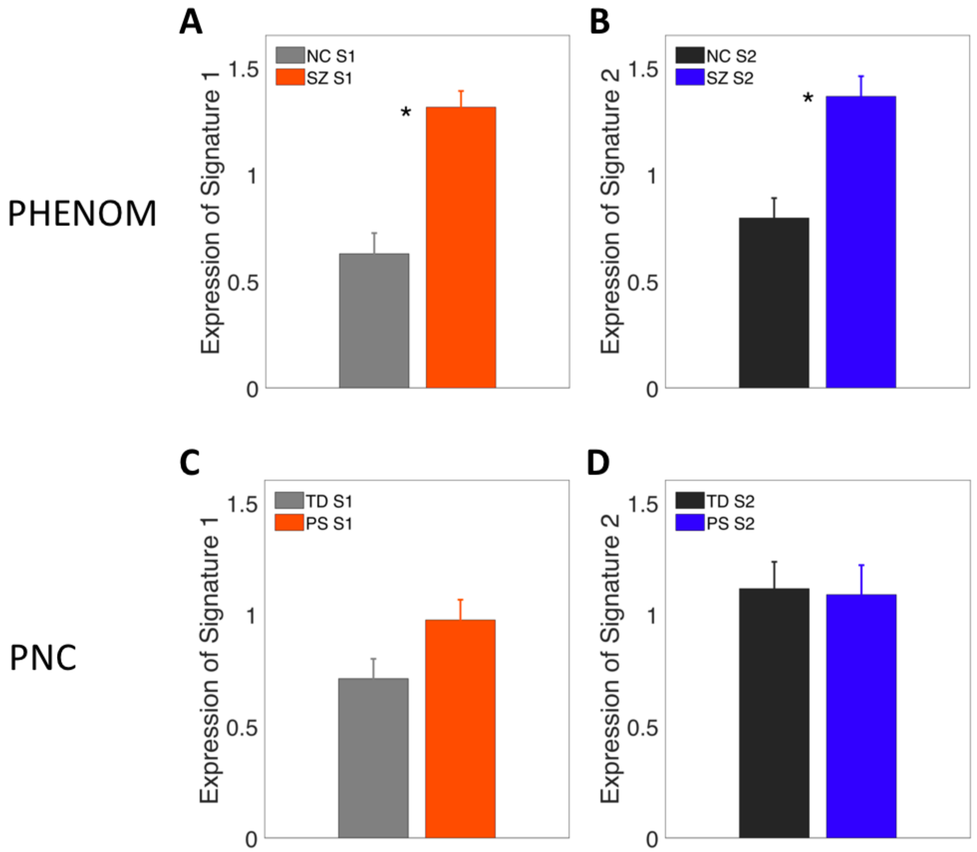


**Figure S7**: Comparison of signature expression strength between TD and PS in PNC, and NC and SZ in PHENOM. In PHENOM, both signature 1 (S1) and signature 2 (S2) are expressed more strongly in SZ than NC (*: $p < 0.05$), but in PNC there was no significant group difference (TD vs. PS) in expression strength for S1 or S2.

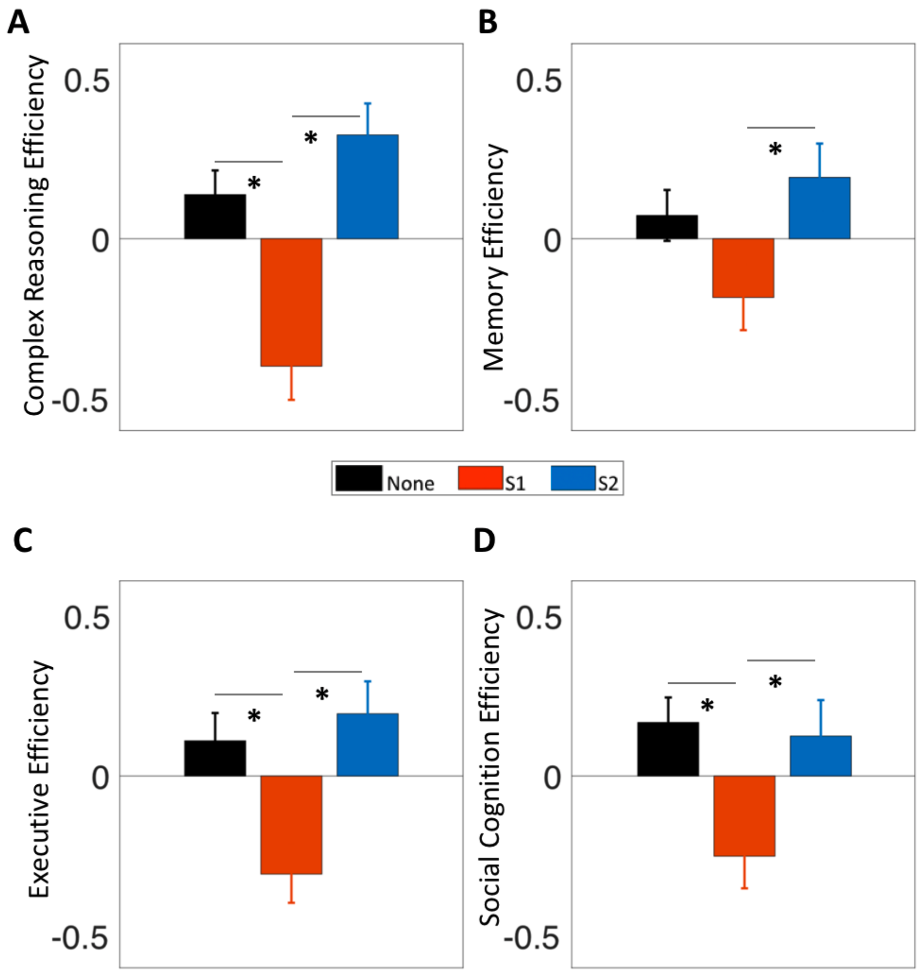


**Figure S8:** Exploratory domain-specific cognitive efficiency comparison between signatures in PNC: Signature 2 (S2) performed significantly better than signature 1 (S1) in complex reasoning, episodic memory, executive, and social cognition domains (*: $p < 0.05$).

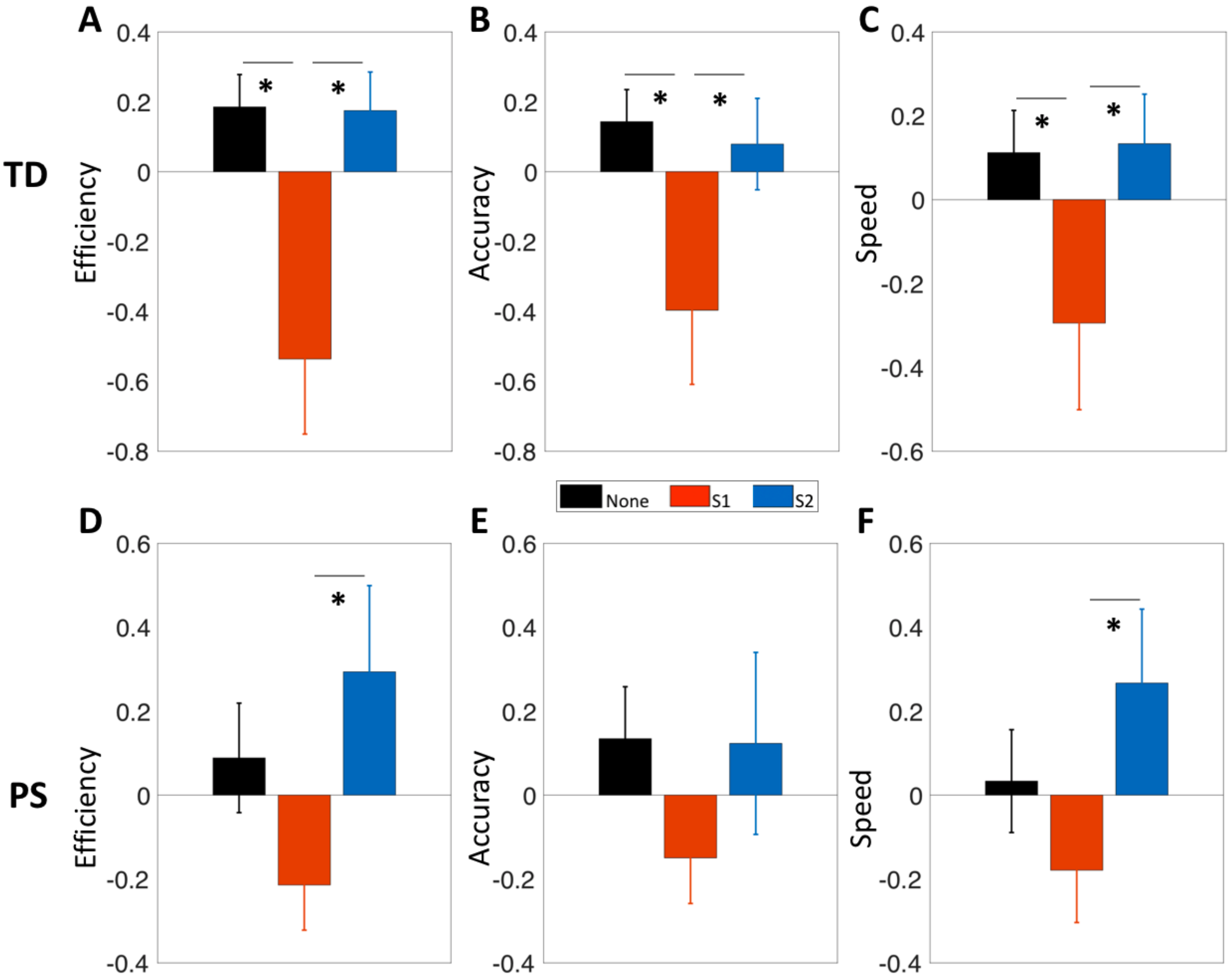


**Figure S9:** Differences in cognitive efficiency, accuracy, and speed between signatures, separately for TD (top) and PS (bottom). In both TD and PS, signature 2 (S2) performed better than signature 1 (S1) (*: $p < 0.05$).

**Table S2:** Clinical and educational profiles between signature 1 (S1) and signature 2 (S2) in PNC (TD) (Note: none of these measures are significant).

| **Variable** | **S1 (N=41)** | **S2 (N=43)** | **S1 vs. S2 *p-value*** |
|---|---|---|---|
| Education, mean years (SD) | 11.73 (1.43) | 11.37 (1.54) | 0.23 |
| Mood, four factors, mean (SD) | -0.11 (1.16) | -0.15 (0.88) | 0.80 |
| Psychosis, four factors, mean (SD) | -0.24 (0.91) | -0.51 (0.84) | 0.86 |
| Externalizing, four factors, mean (SD) | -0.30 (0.76) | -0.16 (0.92) | 0.41 |
| Phobias/fear, four factors, mean (SD) | -0.03 (0.88) | -0.16 (0.80) | 0.44 |
| Overall psychopathology, four factors, mean (SD) | -0.42 (0.83) | -0.34 (0.77) | 0.46 |
| Mood, correlated factors, mean (SD) | -0.41 (0.81) | -0.36 (0.69) | 0.51 |
| Psychosis, correlated factors, mean (SD) | -0.42 (0.79) | -0.38 (0.73) | 0.65 |
| Externalizing, correlated factors, mean (SD) | -0.48 (0.77) | -0.34 (0.79) | 0.43 |
| Phobias/fear, correlated factors, mean (SD) | -0.35 (0.76) | -0.35 (0.74) | 0.99 |
| Overall psychosis, mean (SD) | -0.69 (0.61) | -0.72 (0.60) | 0.75 |
| Positive, two factors, mean (SD) | -0.69 (0.60) | -0.69 (0.54) | 0.84 |
| Negative, two factors, mean (SD) | -0.58 (0.73) | -0.62 (0.68) | 0.86 |
| Positive, three factors, mean (SD) | -0.71 (0.56) | -0.71 (0.53) | 0.83 |
| Positive delusional, three factors, mean (SD) | -0.57 (0.68) | -0.57 (0.58) | 0.68 |
| Negative, three factors, mean (SD) | -0.55 (0.75) | -0.60 (0.70) | 0.99 |
| Psychoactive medication, Yes, N (%) | 5 (12.20%) | 10 (23.26%) | 0.63 |

**Table S3:** Clinical and educational profiles between signature 1 (S1) and signature 2 (S2) in PNC (PS) (Note: none of these measures are significant after multiple comparisons).

| **Variable** | **S1 (N=72)** | **S2 (N=26)** | **S1 vs. S2 *p-value*** |
|---|---|---|---|
| Education, mean years (SD) | 10.54 (1.68) | 10.31 (1.91) | 0.32 |
| Mood, four factors, mean (SD) | 0.19 (1.05) | 0.66 (1.16) | 0.04 |
| Psychosis, four factors, mean (SD) | 0.70 (1.14) | 0.58 (1.00) | 0.64 |
| Externalizing, four factors, mean (SD) | -0.06 (0.89) | 0.17 (0.88) | 0.43 |
| Phobias/fear, four factors, mean (SD) | -0.07 (1.12) | -0.03 (1.09) | 0.95 |
| Overall psychopathology, four factors, mean (SD) | 1.24 (0.56) | 1.19 (0.66) | 0.80 |
| Mood, correlated factors, mean (SD) | 1.12 (0.74) | 1.29 (0.79) | 0.29 |
| Psychosis, correlated factors, mean (SD) | 1.32 (0.53) | 1.23 (0.64) | 0.67 |
| Externalizing, correlated factors, mean (SD) | 0.83 (0.78) | 0.92 (0.59) | 0.49 |
| Phobias/fear, correlated factors, mean (SD) | 0.75 (0.87) | 0.77 (0.81) | 0.79 |
| Overall psychosis, mean (SD) | 1.22 (0.70) | 1.04 (0.81) | 0.52 |
| Positive, two factors, mean (SD) | 1.21 (0.75) | 1.01 (0.91) | 0.54 |
| Negative, two factors, mean (SD) | 1.10 (0.98) | 1.38 (0.99) | 0.18 |
| Positive, three factors, mean (SD) | 1.26 (0.72) | 1.04 (0.87) | 0.32 |
| Positive delusional, three factors, mean (SD) | 0.96 (1.03) | 0.83 (1.10) | 0.69 |
| Negative, three factors, mean (SD) | 1.05 (0.97) | 1.40 (1.15) | 0.09 |
| Psychoactive medication, Yes, N (%) | 12 (16.67%) | 6 (23.08%) | 0.9 |

UKBB

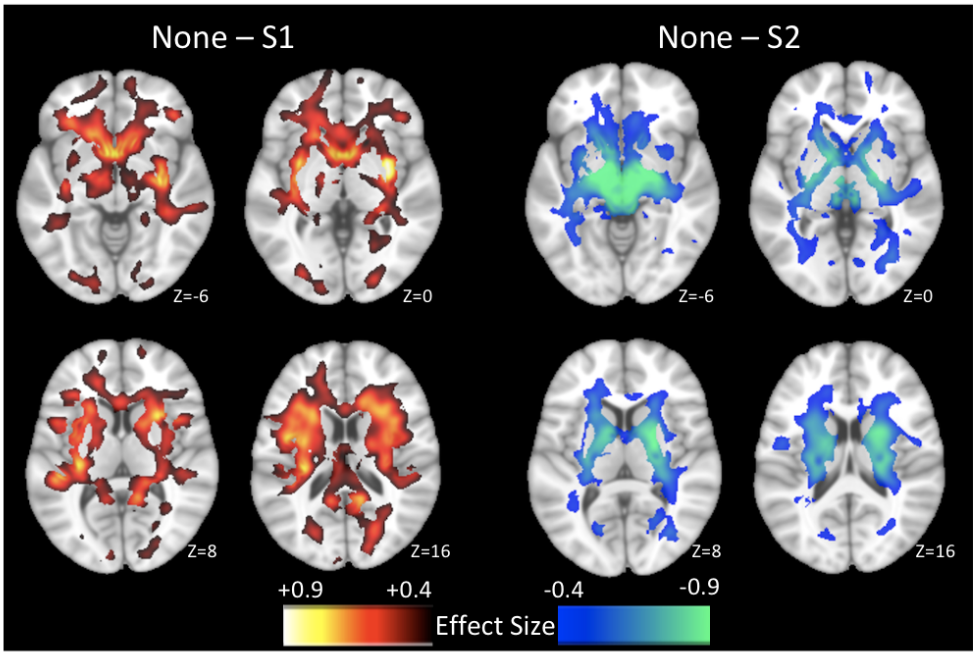


**Figure S10:** Voxel-based comparisons of regional white matter (WM) volumes between individuals expressing primarily signature 1 (S1) or signature 2 (S2), compared to individuals expressing none of these two signatures, in UKBB are displayed for purely visualization purposes. S1 was characterized by widespread reductions of WM volumes, whereas S2 was characterized by increased WM volumes mainly in the internal capsule. (Cohen's d effect size maps were generated by masking MIDAS results after FDR-correction over voxels at $p < 0.05$).

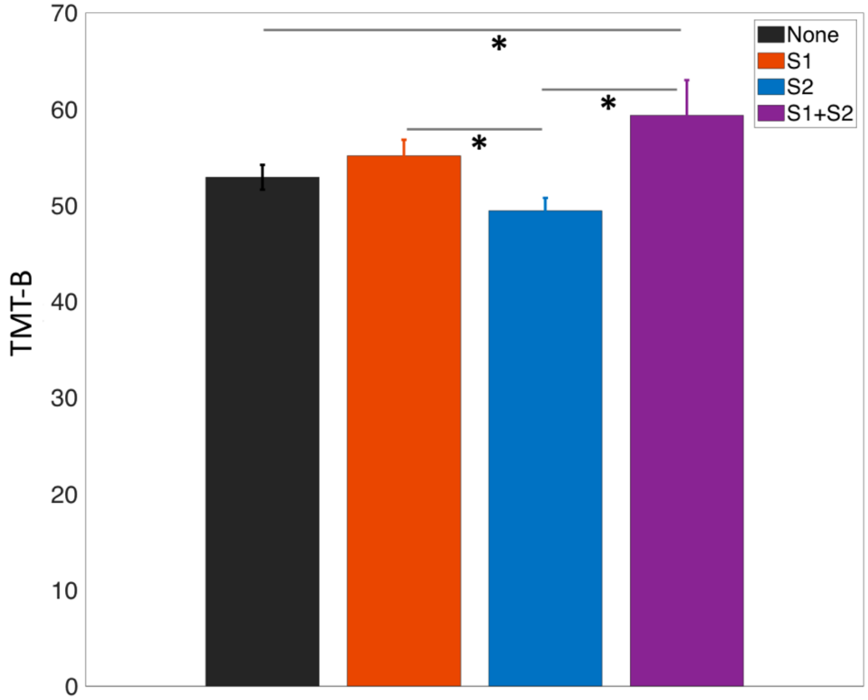

**Figure S11:** Differences in cognitive performance across neuroanatomical signatures in UKBB dataset: participants with schizophrenia signature 2 ('S2') and 'None' took relatively shorter time to complete trail-making test part-B (TMT-B) task than those with schizophrenia signature 1 ('S1') and those with both signatures 'S1+S2' (*: $p < 0.05$).

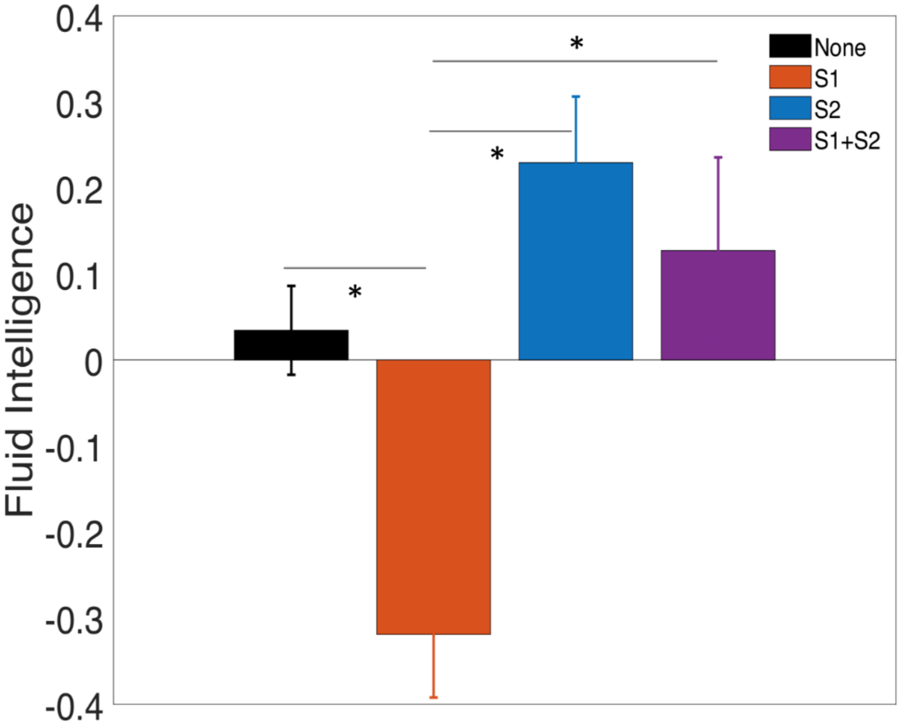


**Figure S12:** Differences in fluid intelligence performance across neuroanatomical signatures in UKBB dataset: participants with schizophrenia signature 1 ('S1') performed worse than those with other signatures (*: $p < 0.05$).

**Table S4:** Clinical, educational, and cognitive profiles between neuroanatomical signature 1 (S1) and signature 2 (S2) in UKBB (N/A: Statistical comparisons were not carried out for measures with small sample size; and note that none of these measures are significant after multiple comparisons correction).

| **Variable** | **S1** (N=203) | **S2** (N=170) | **S1 vs. S2** ***p-value*** |
|---|---|---|---|
| Education, mean years (SD), N | 13.50 (1.58), 198 | 10.36 (1.79), 168 | 0.12 |
| Self-reported depression, Yes (%), N | 11 (8.87%), 124 | 16 (17.20%), 93 | 0.07 |
| Self-reported anxiety, Yes (%), N | 5 (4.03%), 124 | 4 (4.30%),124 | N/A |
| Self-reported nervous breakdown, Yes (%), N | 0 (0%), 124 | 2 (2.15%),93 | N/A |
| Self-reported self-harm, Yes (%), N | 0 (0%), 124 | 1 (1.08%), 93 | N/A |
| Diastolic blood pressure, mean mm Hg (SD), N | 78.84 (10.79), 176 | 77.19 (9.91), 150 | 0.12 |
| Systolic blood pressure, mean mm Hg (SD), N | 129.14 (15.12), 176 | 127.89 (16.35), 150 | 0.29 |
| Current tobacco smoking, Yes (%), N | 28 (13.79%), 203 | 8 (4.71%), 170 | N/A |
| Maternal smoking around birth, Yes (%), N | 54 (30.00%), 180 | 40 (26.49%), 151 | 0.48 |
| Vascular heart problems diagnosed by doctor, Yes (%), N | 14 (6.90%), 203 | 14 (8.24%), 170 | 0.63 |
| Smoking status, Yes (%), N | 75 (37.13%), 202 | 48 (28.24%), 170 | 0.07 |
| Alcohol drinker status, Yes (%), N | 191 (94.09%), 203 | 167 (98.24%), 170 | 0.04 |
| Ever taken cannabis, Yes (%), N | 49 (34.27%), 143 | 48 (40.34%), 119 | 0.31 |
| Body mass index, mean (SD), N | 26.07 (4.67), 202 | 26.52 (4.50), 169 | 0.18 |
| Ever depressed for a whole week, Yes (%), N | 68 (57.63%), 118 | 54 (66.67%), 81 | 0.20 |
| Number of depression episodes, mean (SD), N | 3.24 (3.57), 58 | 8.40 (28.76), 50 | 0.89 |
| Illness injury bereavement stress in last 2 years, Yes (%), N | 92 (45.32%), 203 | 76 (44.97%), 169 | 0.94 |
| Neuroticism score, mean (SD), N | 4.46 (3.43), 202 | 4.55 (3.46), 169 | 0.85 |
| Lifetime number of depressed periods, Yes (%), N | 40 (54.05%), 74 | 33 (56.90%), 58 | 0.74 |
| Ever heard an unreal voice, Yes (%), N | 4 (2.78%), 144 | 5 (4.20%), 119 | N/A |
| Ever believed in an unreal conspiracy against self, Yes (%), N | 2 (1.39%), 144 | 3 (2.52%), 119 | N/A |
| Ever seen an unreal vision, Yes (%), N | 2 (1.40%), 143 | 7 (5.93%), 118 | N/A |
| Ever believed in unreal communications or signs, Yes (%), N | 0 (0%), 144 | 2 (1.68%), 119 | N/A |
| Ever talked to a health professional about unusual or psychotic experiences, Yes (%), N | 3 (50%), 6 | 4 (33.33%), 12 | N/A |
| Ever prescribed a medication for unusual or psychotic experience, Yes (%), N | 2 (33.33%), 6 | 3 (25%), 12 | N/A |
| Digit symbol substitution test, mean (SD), N | 21.72 (0.81), 43 | 22.68 (0.77), 35 | 0.36 |
| Numeric memory, mean (SD), N | 6.90 (0.19), 52 | 7.05 (0.21), 41 | 0.75 |
| Matrix pattern completion, mean (SD), N | 8.39 (0.27), 43 | 8.66 (0.34), 35 | 0.62 |

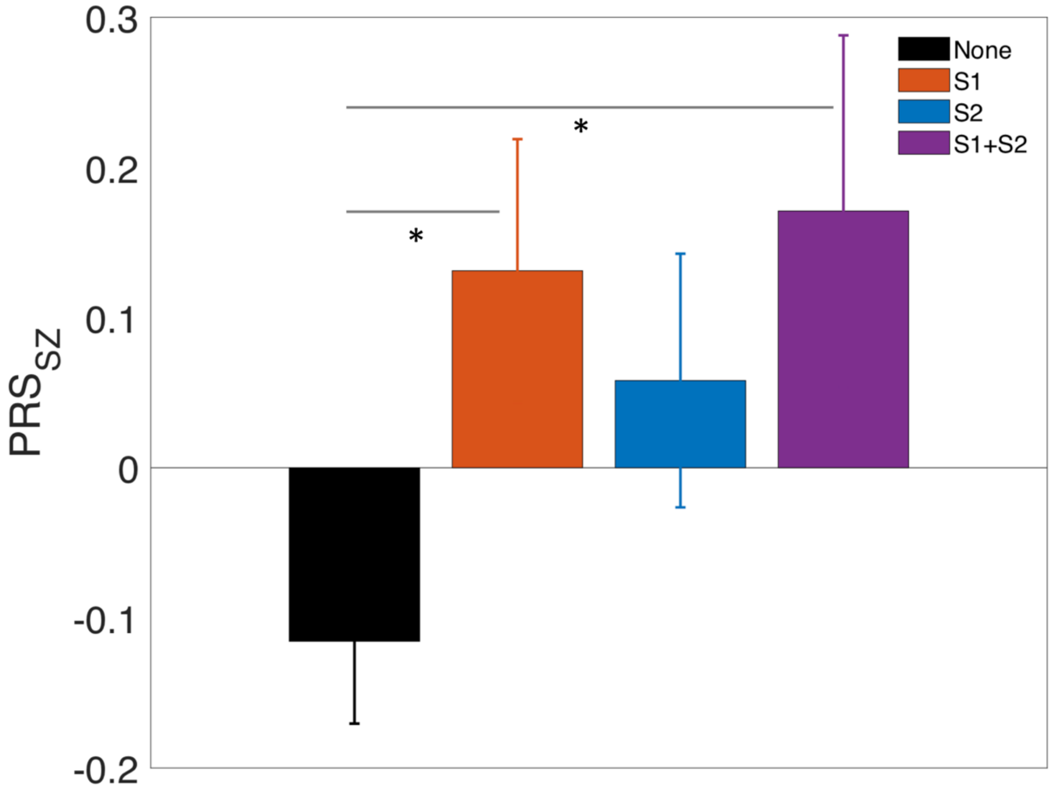

**Figure S13:** Genetic profiles across neuroanatomical signatures in European-ancestry UKBB dataset (*: $p < 0.05$).

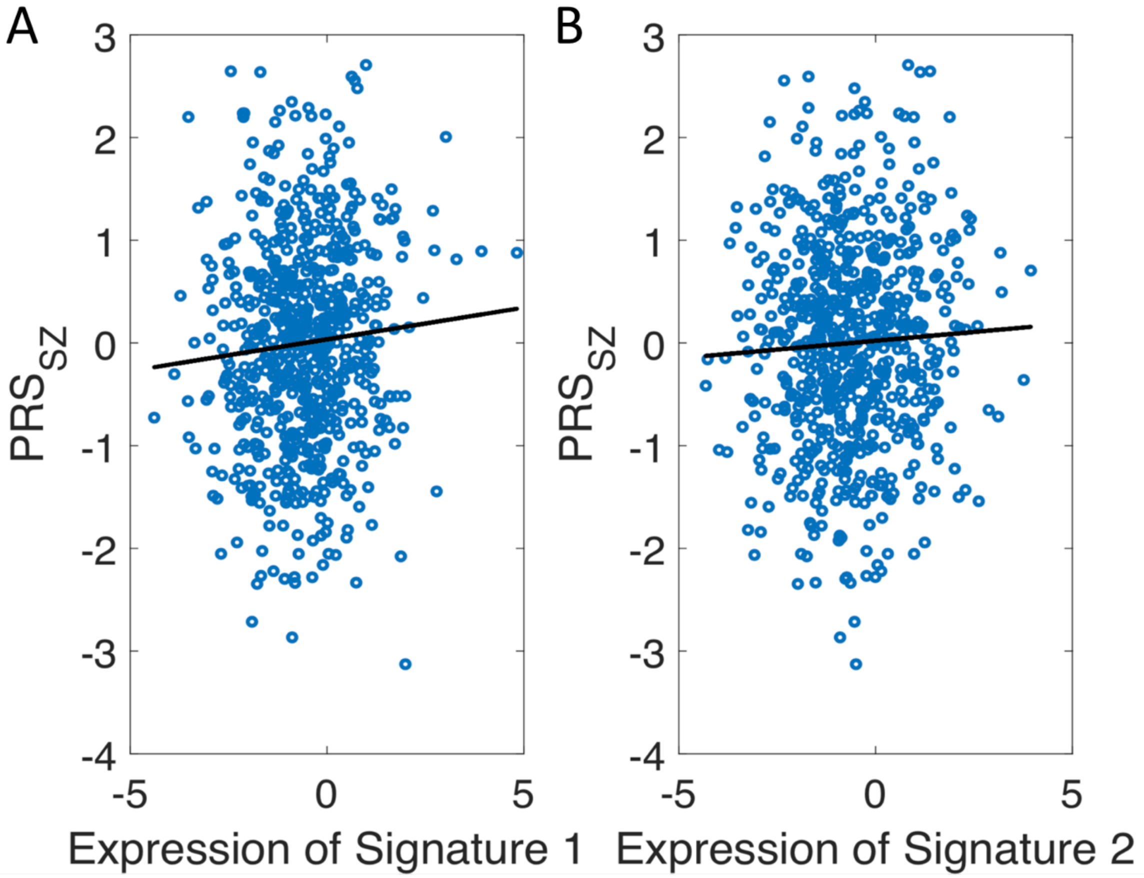


**Figure S14:** Across the full European-ancestry UKBB sample (n = 671), **A**) association between S1 expression and $PRS_{SZ}$ (*ρ = 0.07; p = 0.05*), and **B**) association between S2 expression and $PRS_{SZ}$ (*ρ = 0.04; p = 0.40*).
